# A novel computational methodology for GWAS multi-locus analysis based on graph theory and machine learning

**DOI:** 10.1101/2021.10.22.21265388

**Authors:** Subrata Saha, Himanshu Narayan Singh, Ahmed Soliman, Sanguthevar Rajasekaran

## Abstract

**Background:** Current form of genome-wide association studies (GWAS) is inadequate to accurately explain the genetics of complex traits due to the lack of sufficient statistical power. It explores each variant individually, but current studies show that multiple variants with varying effect sizes actually act in a concerted way to develop a complex disease. To address this issue, we have developed an algorithmic framework that can effectively solve the multi-locus problem in GWAS with a very high level of confidence. Our methodology consists of three novel algorithms based on graph theory and machine learning. It identifies a set of highly discriminating variants that are stable and robust with little (if any) spuriousness. Consequently, likely these variants should be able to interpret missing heritability of a convoluted disease as an entity.

**Results:** To demonstrate the efficacy of our proposed algorithms, we have considered astigmatism case-control GWAS dataset. Astigmatism is a common eye condition that causes blurred vision because of an error in the shape of the cornea. The cause of astigmatism is not entirely known but a sizable inheritability is assumed. Clinical studies show that developmental disorders (such as, autism) and astigmatism co-occur in a statistically significant number of individuals. By performing classical GWAS analysis, we didn’t find any genome-wide statistically significant variants. Conversely, we have identified a set of stable, robust, and highly predictive variants that can together explain the genetics of astigmatism. We have performed a set of biological enrichment analyses based on gene ontology (GO) terms, disease ontology (DO) terms, biological pathways, network of pathways, and so forth to manifest the accuracy and novelty of our findings.

**Conclusions:** Rigorous experimental evaluations show that our proposed methodology can solve GWAS multi-locus problem effectively and efficiently. It can identify signals from the GWAS dataset having small number of samples with a high level of accuracy. We believe that the proposed methodology based on graph theory and machine learning is the most comprehensive one compared to any other machine learning based tools in this domain.

## 1 Introduction

A genome-wide association study (GWAS) is an observational study employed in genetics research to associate a specific set of genetic variations with a particular trait. It is also commonly known as whole genome association study (WGAS) as it considers a genome-wide set of genetic variants. The method skims through the entire genomes from a set of individuals and searches for any variants that can explain the presence of a disease. Genetic variants are of many types, e.g. single-nucleotide polymorphisms (SNPs), insertions and deletions (indels), tandem repeats, copy number variations (CNVs), and so forth. However, GWAS primarily focuses on finding associations between SNPs and traits, such as major human diseases. Nevertheless, it can be applied uniformly to other types of genetic variant as stated above along with other organisms. The variants identified by GWAS now can be used to elucidate how genes harbored by those variants contribute to the disease and potentially develop better prevention and treatment strategies. Please, note that we use the terms variant and SNP interchangeably throughout this article.

One of the foremost weaknesses of GWAS is its inability to deal with the *missing heritability* problem. It is because single genetic variations can explain only a portion of heritability of traits (such as diseases, behaviors, and other phenotypes). Much of the heritability of some complex diseases is missing because a person’s susceptibility to a particular disease may well depend on the combined effect of a set of variants. Missing heritability problem is presumed to be partly rooted in the inherent weakness of the classical statistical methods followed by GWAS. Traditional statistical methods [1] are designed to analyze susceptibility of variants in GWAS by considering only a single variant at a time. On the contrary, it is proven that multiple variants act together to cause many common diseases. These complex interactions among variants are known as multi-locus interactions [2]. Contemporary case-control studies fail to identify multi-locus effects by using the traditional *p*-value calculations and a large amount of potentially available information is lost (24). There are numerous challenges in designing and analyzing the joint effects of multiple genetic factors. For example, in a typical GWAS genotypes of up to 100 million variants are sequenced from not more than several thousands of individuals. Here we have many more variables *p* than samples *n*. Classical models require *p* < *n*, whereas in GWAS we have *p* ≫ *n*. We also need to account for correlations, such as linkage disequilibrium (LD) between variants. Therefore, standard multi-variable statistical approaches like multiple linear or logistic regression are not very promising tools to detect complex multi-locus interactions from genome-wide data. Fortunately, machine learning algorithms provide several intuitive alternatives to perform multi-locus analyses within acceptable time, accuracy, memory, and money.

Two types of analytic methods have been developed to elucidate GWAS datasets. Some methods infer phenotypic risk of an individual based on the given genetic information while the rest focus on identifying statistically significant variants that can explain the given trait. In this proposed research we intend to combine both objectives by developing a robust and stable multi-locus association methodology that can identify a subset of discriminating variants that can together explain a complex disease as-well-as predict a person’s susceptibility to that disease. In this context, we have designed and developed three interconnected algorithms to decipher the biology of complex diseases by employing GWAS case-control datasets. Some of the main objectives of this work include but not limited to: (1) not to use any type of raw *p*-value threshold; (2) reduce variants with minimal information loss; (3) identify a subset of discriminating variants that are stable and robust across multiple bootstrapped samples; and (4) instrument a theoretical framework to assess the robustness and statistical significance of a machine learning model of interest.

There exist several machine learning (ML) based methods to solve multi-locus problem in GWAS [3, 4, 5, 6, 7, 8]. An excellent comparative study of several machine learning methods with respect to GWAS can be found in [9]. All the methods cited above applied ML tools directly on the reduced set of variants by imposing certain kinds of threshold, such as *p*-value threshold. Like any other statistical methods, ML algorithms also suffer from very underdetermined systems where we have many more variables compared to the number of samples. Fortunately, ensemble techniques [10] can be employed in the context of multi-locus problem in GWAS to elucidate hidden and complex biology of a compound disease. To the best of our knowledge, there exists no comprehensive ML based computational methodology in the current literature of GWAS for the identification of a robust and stable set of discriminating variants that can potentially explain the biology of an underlying disease. To demonstrate the relevance and effectiveness of our proposed methodology, we have employed it on a case-control GWAS dataset where cases consist of individuals having astigmatism (an imperfection in the curvature of eye’s cornea or lens). Controls do not have any type of astigmatisms. Individually, each SNP in this dataset has very limited predictive power. For example, after performing proper quality control procedures, the smallest raw *p*-value as shown in Figure 1 is not genome-wide significant (Benjamini-Hochberg adjusted *p*-value is 0.97). Therefore, classical GWAS analysis fails to identify any discriminating variants from this particular dataset. On the contrary, our methodology identifies 350 robust and stable variants collectively having a high discriminating power (diagnostic odds ratio is 3.25 in validation samples). Please, note that some of these variants have very large *p*-values (for instance 0.3, 0.4, etc.). It indicates that applying a *p*-value threshold can have an undesired effect on any type of multi-locus genetic analysis.

**Figure 1:**
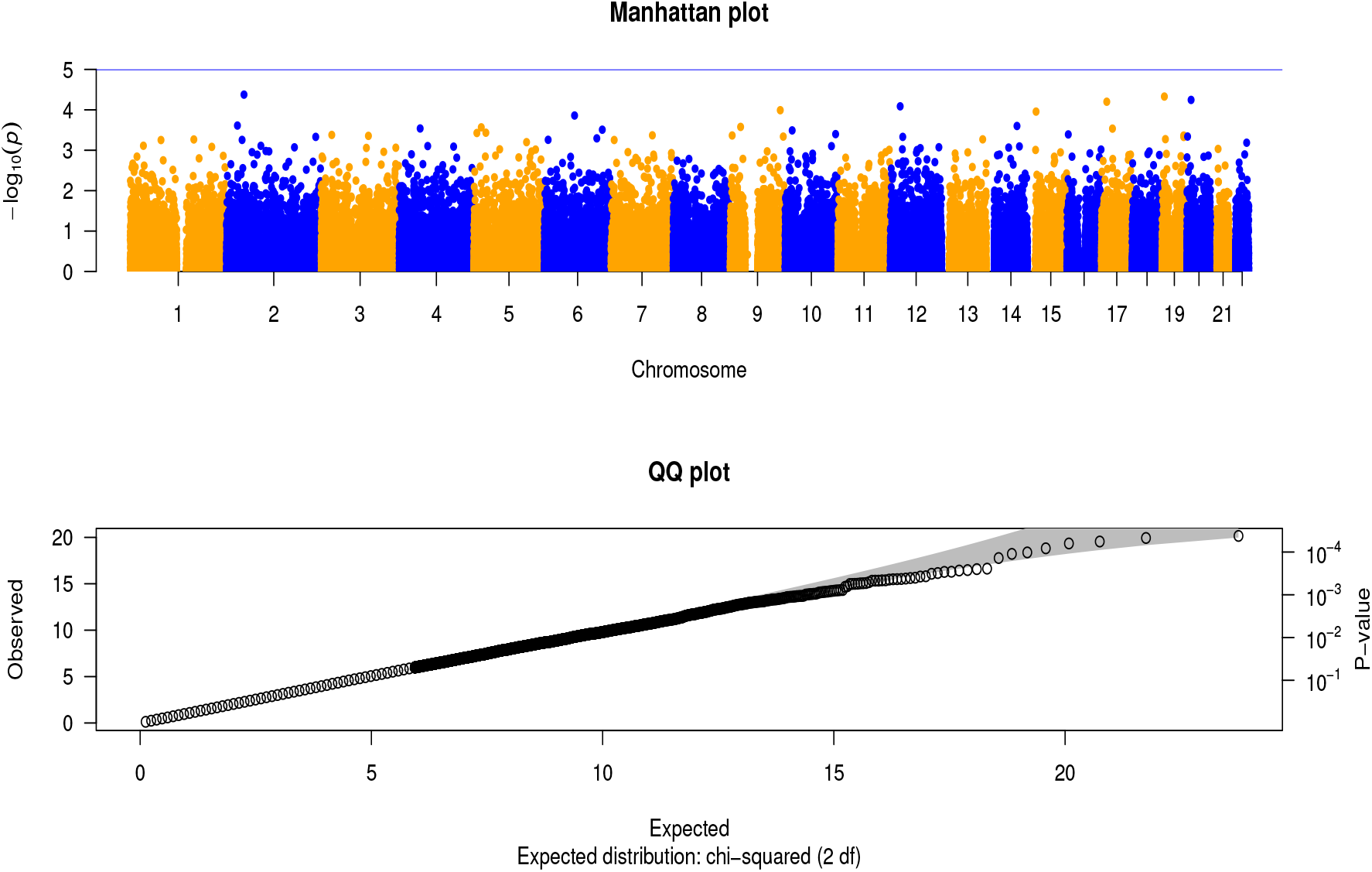
*p*-values from logistic regression based on entire astigmatism GWAS dataset.

## 2 Methods

Our computational methodology consists of three interconnected algorithms to accomplish three fundamental objectives. At the beginning, we retain unique variants and discard duplicate ones in terms of their values across the samples. We then select a set of representative variants using linkage disequilibrium measures. Finally, a subset of highly discriminating variants is identified using a robust and stable machine learning algorithm. Next, we illustrate each step in detail.

### 2.1 Selecting unique SNPs

In a typical GWAS, we have roughly 10-100 million variants. Consequently, there is a high chance that multiple variants may have identical values across the samples. There will be no information loss if we discard the duplicates. However, we have an enormous number of variants as stated above. In this context, we have developed a randomized algorithm that can exactly identify the duplicates using random sampling and bitwise operations. It works as follows. Traditionally, biallelic variants are encoded with 0, 1 or 2 numerals based on the number of copies of non-reference alleles, e.g. 0 refers to the homozygous with the reference allele, 1 means heterozygous with the reference allele, and 2 indicates homozygous with the alternate or mutant allele. Typically, a character is represented by 8 bits in ASCII (American Standard Code for Information Interchange) format. We can encode each biallelic variant using 2 bits (i.e. 0, 1, and 2 can be represented by 00, 01, and 10, respectively). As a result, the dataset will be reduced by 4-*fold* using 2-bit encoding. It will also provide us the opportunity to compute Hamming distance between any pair of variants using bitwise operations that is tremendously faster than the string-matching operation.

After encoding each variant across all the subjects, we randomly sample a subset of the coordinates in the encoded binary space and hash the variants based on this subset. Two variants will be hashed into the same hash bucket if they have identical values for the randomly chosen coordinates (i.e., subjects). If two variants fall into the same bucket in the hash table, this is a candidate pair. For each bucket, we compute the Hamming distance between each pair of variants by considering all the coordinates using bitwise operations (i.e. XORing and then counting the number of 1s in the binary space; if the number of 1s is zero, Hamming distance will be zero). We keep a graph where each variant acts as a node. Two nodes will be connected by an edge if the Hamming distance between them is zero. At the end, the graph contains a set of connected components (strictly speaking, each connected component will be a clique). We then randomly select a variant from each of the connected components. These selected variants will be unique across the entire space of variants. The details of the algorithm can be found in Algorithm 1.

### 2.2 Selecting representative variants

In population genetics, linkage disequilibrium (LD, in short) is the non-random association of alleles at different loci in a given population. Loci are said to be in linkage disequilibrium when the frequency of association of their different alleles is higher or lower than what would be expected if the loci were independent and associated randomly [11]. Consider two biallelic loci, locus 1 with alleles *a* and *A* and locus 2 with alleles *b* and *B*. Suppose the frequencies for alleles *a* and *A* are *p*_*a*_ and 1 − *p*_*a*_, respectively and the frequencies for alleles *b* and *B* are *p*_*b*_ and 1 − *p*_*b*_, respectively. The *r*^2^ measure of linkage disequilibrium is defined as:

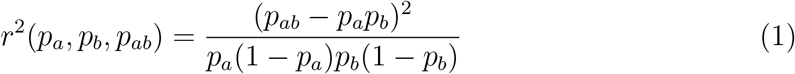

where *p*_*ab*_ is the frequency of haplotypes having allele *a* at locus 1 and allele *b* at locus 2 [12]. As the square of a correlation coefficient, *r*^2^(*p*_*a*_, *p*_*b*_, *p*_*ab*_) can range from 0 to 1 as *p*_*a*_, *p*_*b*_ and *p*_*ab*_ vary. If *r*^2^ ≈0 at two different loci *x* and *y*, they are said to be in linkage equilibrium, i.e., those two loci *x* and *y* are uncorrelated. Conversely if *r*^2^ ≈1, they are in linkage disequilibrium. In such a scenario we can conclude that *x* and *y* are highly correlated and so, removing one would cost minimal loss of information.

To effectively prune the set of correlated variants based on LD measures, we have designed and developed a novel graph theoretic algorithm. It works as follows. Given that we have a set of variants *S* and pair-wise LDs among the variants (please, note that 400kb window size was used while computing pair-wise LDs across the chromosomes), we create a graph *G*(*V, E*) where each variant *s ∈ S* is treated as a node *n*. Two nodes *n*_1_ and *n*_2_ in graph *G* are connected by an edge *e* if *LD*(*n*_1_, *n*_2_) ≥ *λ*, a user defined threshold (*λ* = 0.9 in our experiment). After constructing such a graph *G*, we compute the centrality score of each node *n ∈ G* using PageRank algorithm. It gives each node in the graph a rating of its importance recursively so that an important node will get more score if it is connected to other important nodes [13].

Let *n*′ be a node with the highest centrality score across the network. We delete each neighboring node *n″* of *n*′ including *n*′ from *G*. We record *n*′ as a representative of its neighbors *n*″. Since, all the nodes *n*″ are in high LD with *n*′, they are highly correlated with *n*′. So, deleting neighboring nodes *n*″ will reduce the dimension of the variant space without any serious loss of information. The same procedure is repeated until all the nodes *n ∈ G* are isolated, i.e., there is no edge *e* in *G*. We record all such nodes *n* as representatives. The details of the algorithm can be found in Algorithm 2.

### 2.3 Selecting a subset of high discriminating variants

To effectively identify a subset of discriminating variants that can elucidate a phenotype (such as a disease), we have developed a robust and stable feature selection algorithm by cleverly assembling a set of linear SVMs (LSVMs) [14, 15, 16]. It works as follows. Assume that we have a set of LSVMs *C* and a GWAS dataset *D. D* is essentially a matrix *M* × *N* where *M* and *N* refer to the number of samples and variants, respectively. Each variant is encoded by 3 numerical values (e.g. 0, 1, and 2) as stated earlier. At the very beginning, we bootstrapped dataset *D* by randomly choosing samples with replacement to form a marginally distinct dataset *D*′. Each LSVM is trained with a slightly different dataset *D*″ by randomly choosing *p*% (= 90% in our experiment) of samples with replacement from dataset *D*′. Consequently, we will have slightly different weights for each of the variants for different LSVMs. Please, note that the weight produced by LSVM for a particular variant is directly proportional to its importance, i.e., the more the weight of a variant, the more will it be significant. We sort the normalized weights of each variant and take the average across the middle 50% weights. I.e. we consider the interquartile range (IQR) of weights while averaging them. It is useful in discarding the outliers. The variants are then sorted in decreasing order based on their average weights. To find a subset of discriminating variants, we linearly search the sorted variants by considering top 10, top 20, top 30, …, top *N* variants. For each of the top set of variants, we compute Matthews correlation coefficient (MCC) using 10-*fold* cross-validation (https://en.wikipedia.org/wiki/Matthews_correlation_coefficient). MCC is mathematically formulated as follows:

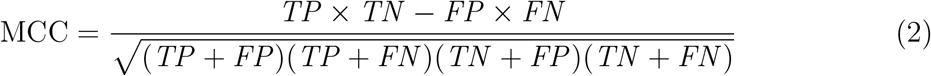

In this above equation, *TP, FN, FP* and *TN* are the number of true positives, false negatives, false positives, and true negatives, respectively. It is a measure of the quality of a binary classifier. Now, from all the set of top variants, we retain the top set having the highest MCC.

The above procedure is iterated multiple times and in each iteration, we get a set of top variants *S*_*i*_. The variants are then sorted based on the number of times it is found in those sets *S*_*i*_. The ordered variants are then searched by taking top 1, top 2, top 3, … variants and for each top set we compute MCC using 10-*fold* cross validation as stated earlier. Finally, we return the top variants having highest MCC. The details of the algorithm can be found in Algorithm 3.

#### Algorithm 1 Algorithm for selecting unique SNPs

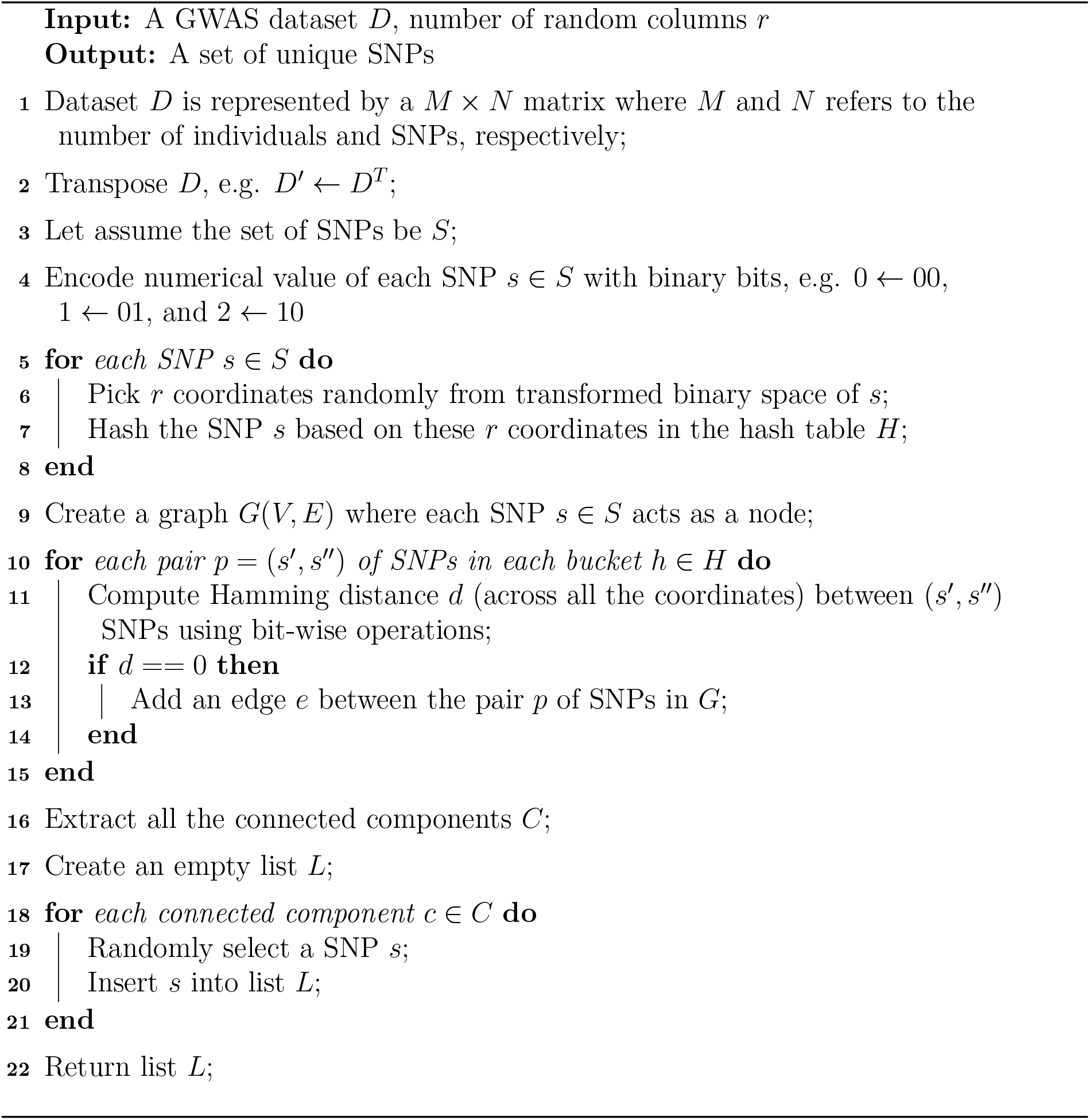

### 2.4 Analyses of our methods

Let the subjects be labelled 1, 2, …, *M* and let the SNPs be labelled 1, 2, *…, N*. Also, let 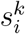 denote the value of SNP *k* for subject *i*, 1 ≤ *I* ≤ *M*; 1 ≤ *k* ≤ *N*.

Since any SNP value is in the set {0, 1, 2}, it follows that the probability that SNPs *i* and *j* (for any two *i* and *j*) have identical values for all the subjects is 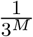 under the uniform distribution model. This also means that the expected number of pairs of SNPs that are identical is 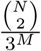

#### Algorithm 2 Algorithm for selecting representative SNPs

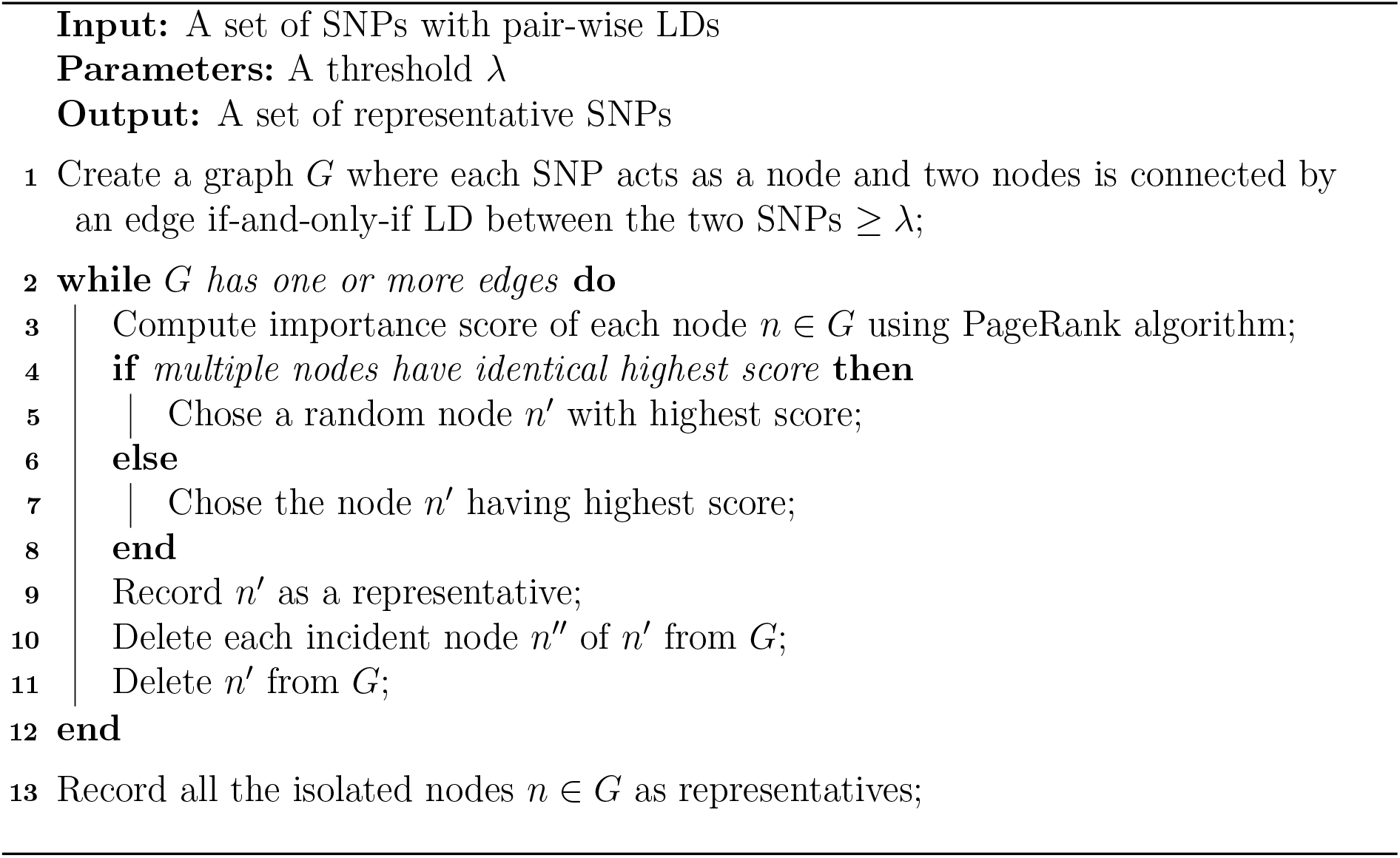

#### Algorithm 1

In Algorithm 1, we perform hashing based on randomly picked *q* subjects (for some suitable value of *q*). This hashing takes *O*(*Nq*) time. If two SNPs are identical across all the subjects, then, clearly these two SNPs will be hashed into the same bucket. Under the assumption that each SNP for each subject has been picked uniformly randomly, we would expect the size of each bucket to be 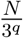.

Within each bucket we compute the Hamming distance between each pair. This will take an expected 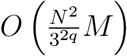 time per bucket. After this step, constructing a graph and identifying the connected components will take an expected 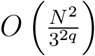 time for each bucket.

Thus the total expected run time is 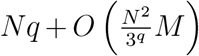 If *q* is chosen to be log_3_ *N*, then this expected run time becomes *O*(*MN*) (assuming that *M* ≥ log_3_ *N*). In this case, note that, this algorithm is asymptotically optimal in expectation.

An alternative to constructing a graph is to employ integer sorting. After hashing the SNPs, we can sort the SNPs within each bucket using integer sorting (with *O*(1) bits at a time). The expected sorting time per bucket will be 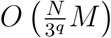. Summed over all the (3^*q*^) buckets, the total run time will be *O*(*MN*).

#### Algorithm 2

In Algorithm 2, the time needed to compute the *r*^2^ linkage disequilibrium (LD) value between any two SNPs is *O*(*M*). Thus the time needed to compute the LD values for every pair of SNPs is *O*(*N*^2^*M*). Followed by this, the graph *G*(*V, E*) can be constructed in *O*(*N*^2^) time. Subsequently, we have a **while** loop in lines 2 to 12. In each iteration of this loop, we compute the importance scores of the nodes. This will take *O(*|*E* |) time. If there are *k* iterations of this **while** loop, then the run time of this loop will be *O*(*E k*). In the worst case, *k* could be *N −* 1. Thus the worst case run time of the **while** loop is *O*(| *V* | | *E*|) = *O*(*N* ^3^).

Put together, the overall run time of Algorithm 2 is *O*(*N* ^3^+*N*^2^*M*) = *O*(*N* ^3^). However, in practice, the value of *k* is small and hence the algorithm runs really fast. For instance, if *k* = *O*(1), then the run time of Algorithm 2 will be *O*(*N*^2^*M*).

##### Algorithm 3 Algorithm for selecting subset of discriminating SNPs

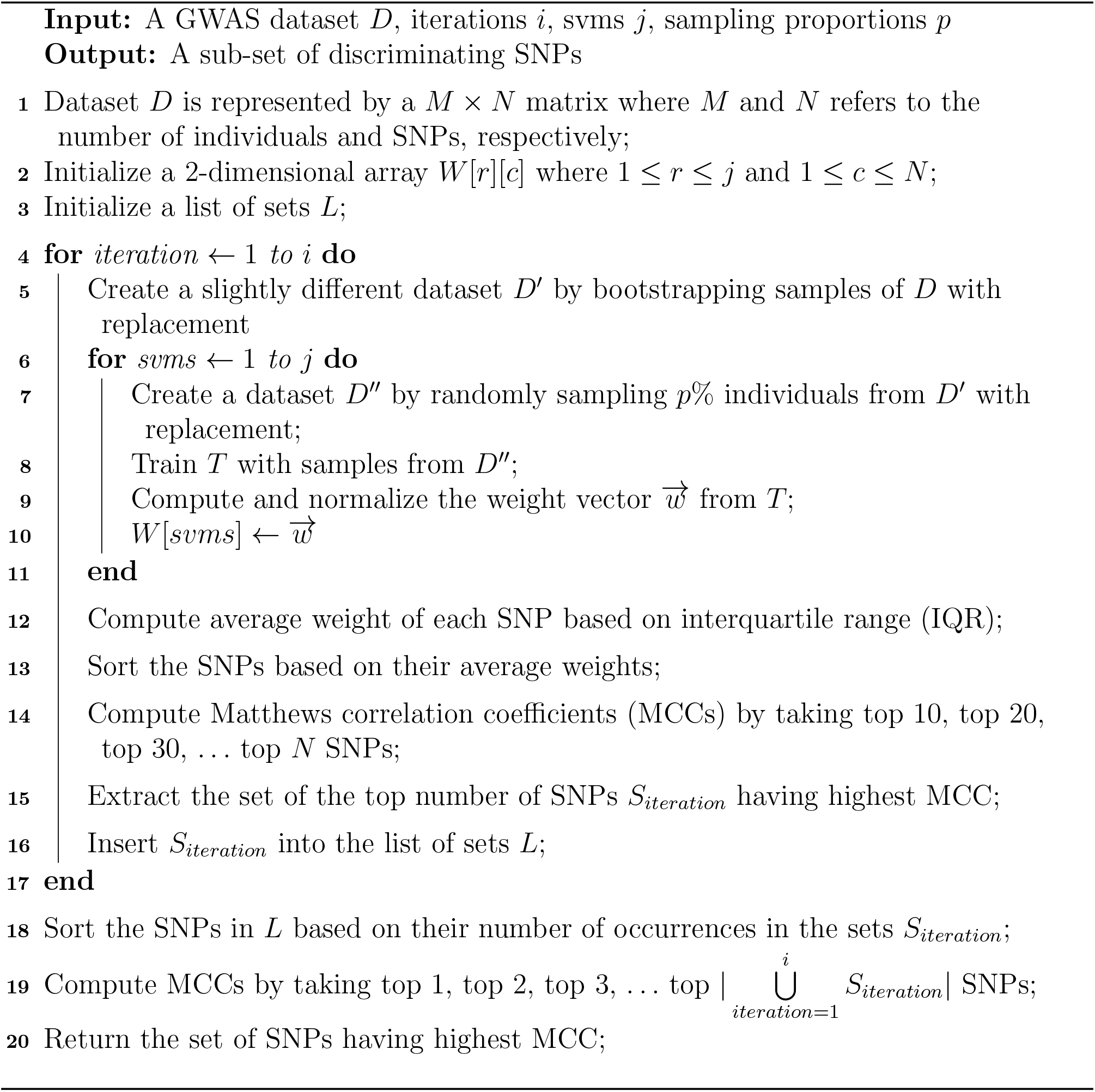

#### Algorithm 3

One of the algorithms repeatedly used in Algorithm 3 is that of training a linear SVM. The size of the training data is *pM* where *p* is the sampling rate (input as a fraction by the user). There are *N* features. A generic algorithm will have a run time of *O*((*pM*)^3^*N*). However, there are faster algorithms. For example, the algorithm of Joachim [17] takes *O*(*pMN*) time for classification problems and *O*(*NpM* log(*pM*)) time for ordinal regression problems.

To keep our analysis generic, let *L*(*m, n*) stand for the time needed to train a linear SVM when there are *m* samples and the number of features is *n*.

The **for** loop of line 4 takes time *O*(*j T* (*pM, N*)). Lines 12 to 16 take a total of *O*(*N* log *N*) time (for one execution). As a result, the run time of the **for** loop of line 4 is *O*(*i*(*j T* (*pM, N*) + *N* log *N*)).

Lines 18 to 20 take a total of *O*(*N* log *N*) time.

In summary, the total run time of Algorithm 3 is *O*(*i j T* (*pM, N*) + *i N* log *N*).

### 2.5 Tools used for biological analyses

Pathogenicity of each of the disseminating variants was predicted using Ensemble [18], which employed various algorithms, namely SIFT [19], PolyPhen [20], CADD [21], REVEL [22], MetaLR [23], Mutation Assessor [24], among others. We subsequently checked whether these associations were related to Astigmatism or related diseases using PheGenI Phenotype-Genotype Integrator (September 2021) [23]. The conserved regulatory regions (cCREs: candidate cis-Regulatory Elements) were extracted from the UCSC genome browser for GRCh38 assembly which was further converted to GRCh37 assembly using LiftOver (Version 377) [25]. Distance between the variants and cCREs were obtained using the BEDTools (version 2.30) [26]. *RegulomeDB* was used to find the regulatory potential of the discriminating variants [27].

## 3 Results

To demonstrate the reliability of our algorithm, we have performed rigorous experimental evaluations on a GWAS dataset consisting of 2 contrasting sets of individuals, i.e. cases and controls. Cases constitute individuals with known astigmatism where controls refer to the healthy individuals with no traces of astigmatism. Note that astigmatism is a common vision problem. It is caused by an error in the shape of the cornea or lens. It can make our vision blurry, fuzzy, or distorted. The causes of astigmatism are not conclusive, but according to various findings it is believed that the genetics is a big factor.

### 3.1 Dataset

The cohort for our study has been constructed by taking variants from *OpenSNP* [28] *using an R package known as rsnps* (version 0.4.0) [29]. Through this software tool, we have downloaded the genetic variation data by carefully choosing each individual’s reported phenotypes to build our case-control study dataset. In this scenario, we were only interested in participants with reported *astigmatism* phenotype. If the reported *astigmatism* phenotype of an individual is any one of the following terms, e.g. “True”, “Mild”, “Right eye only”, “Left eye only”, “Extreme, bilaterally”, “Yes - very minor”, and “Severe”, we include the subject in the case group. On the other hand, if the reported *astigmatism* phenotype of a person is any one of these terms, e.g. “No astigmatism” and “False”, we include him/her in the control group. Please note that some of the individuals were discarded from our study because of data format errors, availability of acceptable genetic data, and unzipping failures. In summary, we enlist 388 and 127 individuals with astigmatism and no-known-astigmatism, respectively.

After selecting the individuals for our study, we extract their genotype data and form *ped* and *map* files suitable for *plink* software tool [30, 31]. We followed proper quality control (QC) measures as described in [32]. Note that we retained only those variants having MAF ≥ 5% as well as genotype missingness < 1%. After performing proper QC along with MAF and missingness cutoffs, the final dataset to be fed into our algorithms consists of 218,328 bi-allelic variants, 311 cases, and 103 controls with a genotyping rate of 0.997848.

### 3.2 Evaluation metrics

#### 3.2.1 Classification accuracy

To assess the performance of our proposed methodology, we have divided our study dataset into two disjoint and unequal parts (i.e. training and validation) by randomly choosing individuals from our study dataset without replacement. Training cohort consists of 60% samples and validation cohort contains the rest 40% samples. The proposed framework was executed only on training dataset. The set of discriminating variants identified by our algorithms is then employed to build a classification model by using the training samples. The model is then employed to predict the class label of each individual in validation cohort. Since, we know the actual class labels of individuals in validation cohort *a priori*, we can now build a confusion matrix as well as can compute various types of performance scores to evaluate our training model, such as diagnostic odds ratio (DOR) (https://en.wikipedia.org/wiki/Diagnostic_odds_ratio). It is defined as the ratio of the odds that the patient tests positive with respect to the odds of testing positive being healthy. DOR is mathematically formulated using the confusion matrix as follows:

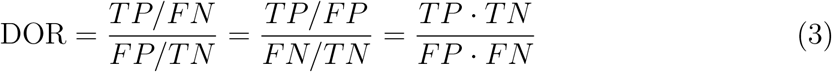

It is extensively used in the domain of medical testing in binary classification that does not depend on the prevalence of the disease. The added advantage is the possibility of constructing its confidence interval. However, we will get only one such data point from a specific scoring criterion and so, we wouldn’t be able to measure the robustness along with statistical significance of our classifier.

To mitigate the issues stated above, we develop an innovative idea as discussed below. Assume that the training cohort is *T* and the set of discriminating variants identified by our framework is *S*. We filter out all the variants from *T* except *S* and form a tailored dataset *T*′ for model construction. Now, we subsample *T*′ (e.g. retaining 90% of total samples in *T*′ without replacement) to construct a predictive model *M*. Subsequently, we compute the DOR of *M* by predicting the class label of each sample from the validation cohort. We repeatedly perform the above procedure multiple times and record the corresponding DOR. These DORs are then transformed to log of DORs (LDOR). As LDOR is normally distributed, we are now able to compute the statistical significance of our training model along with a confidence interval.

#### 3.2.2 Biological significance

The biological significance analysis is based on various biological models, e.g. variant effect prediction, variant-disease association, disease-gene association, gene ontology (GO) terms, disease ontology (DO) terms, and biological pathway enrichment and network analyses. Please note that GO and DO terms analyses were performed using *cluster-Profiler* [33]. *Biological pathways are extracted from ConsensusPathDB-human* (CPDB) database [34]. The functional consequences as well as the effect of our identified variants are analyses through Ensembl Variant Effect Predictor (VEP) [35].

### 3.3 Outcome

The cause of astigmatism is inconclusive. It is widely accepted that genetic factors might have some influences on it [36, 37]. According to American Academy of Ophthalmology (AAO), astigmatism is caused by an irregular curvature of the eye’s cornea or lens and the likelihood of developing astigmatism is inherited (https://www.aao.org/eye-health/diseases/what-is-astigmatism). It can be developed after an individual suffered from an eye disease, eye injury, or surgery. In Europe and Asia, astigmatism affects 30% to 60% of adults [38]. People of all ages and ethnicity can be affected by astigmatism. In this experimental evaluation, we wanted to validate as well as extend some of the observations and hypotheses established in this domain.

#### 3.3.1 Variant analysis

After executing our algorithmic framework on around 218k SNPs, we obtained a set of 350 highly discriminating variants, say *S*. We build a learning model based on *S* using only the training examples. The model is then assessed using the validation examples and the DOR produced by our model is equal to 3.25 and the 95% confidence interval is (1.12, 9.44). To assess the robustness as well as statistical significance of our proposed algorithms, we employ our theoretical framework as described in *Evaluation metrics* subsection. As evident from Figure 2, it is nicely fitted with various theoretical distributions. The average LDOR is 0.96 and the corresponding 95% confidence interval lies in (0.96, 1.02) based on *t*-distribution. We state the null hypothesis as the learning model doesn’t have any predictive power (i.e., the mean of LDOR is zero). The *t* score is 64.89 and the corresponding 2-tailed *p* value is 0 and so, we reject our null hypothesis. Accordingly, the learning model delivered by our algorithms is highly predictive and the variants identified should together elucidate the underlying complex biological system of astigmatism.

**Figure 2:**
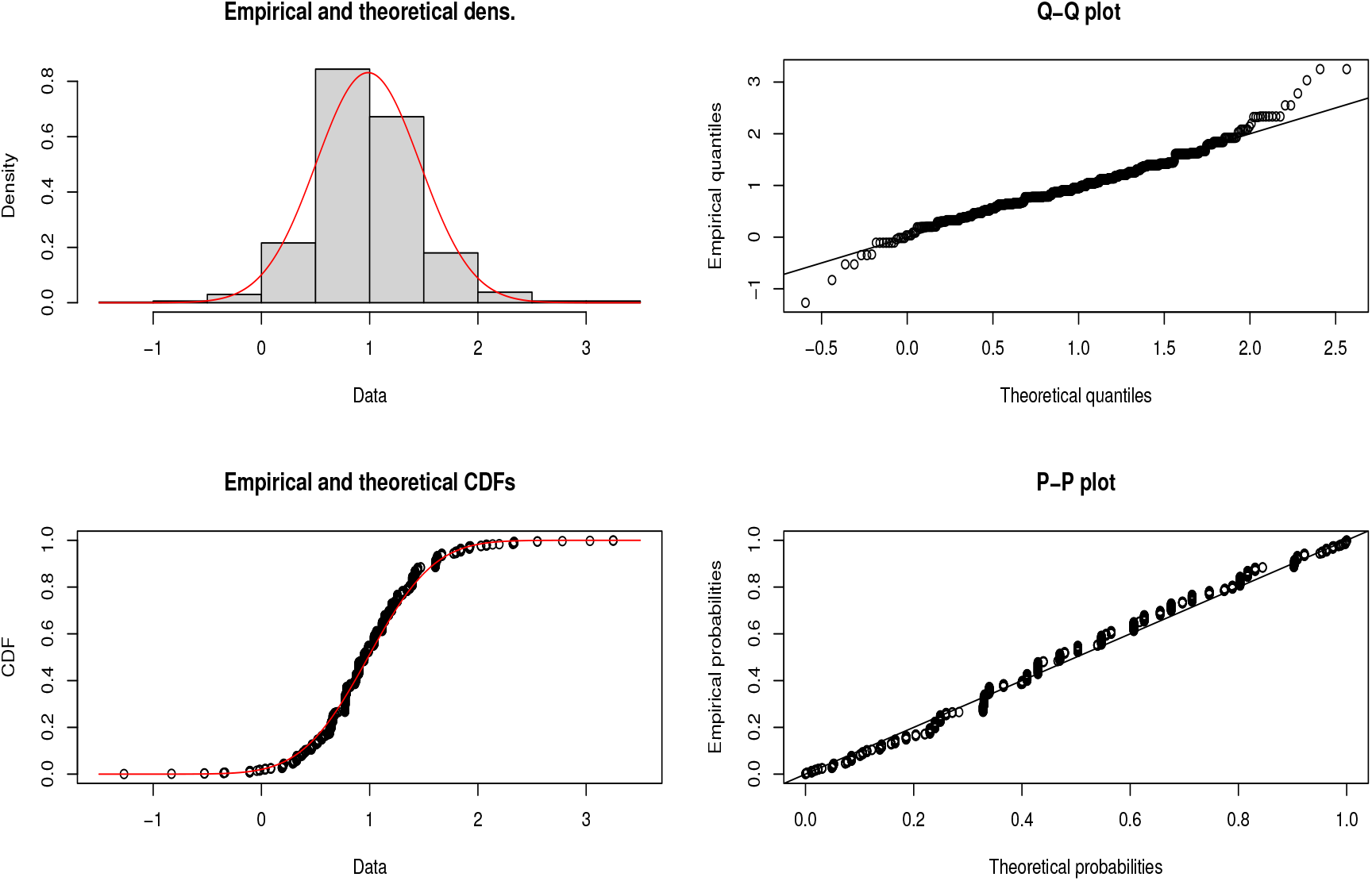
Assessment of validation examples using the model built from training examples.

A set of variants *S*′ (detected by our Algorithms 1 and 2) that were identical and/or in high linkage disequilibrium with some of the variants in *S*, we add them with *S* to get *S*″(= *S* ⋃ *S*′) where |*S*″ | = 379. We employed ensemble variant effect predictor (VEP) to determine the effect of our variants *S*″. Figure 3 shows the relative abundances of the functional consequence of *S*″. 7 SNPs (around 2%) are annotated as *deleterious* (SIFT score ≤ 0.05), i.e. these variants are known to affect their proteins’ function and possibly contribute to genetic diseases. The genes harbored by those variants can be found in Table 1.

**Table 1:** Genes harbored by deleterious variants identified by our method.

| Variant | Gene symbol | Gene ID | Gene name |
| --- | --- | --- | --- |
| rs13212023 | ADGRF2 | 222611 | adhesion G protein-coupled receptor F2 |
| rs2769058 | C9orf40 | 55071 | chromosome 9 open reading frame 40 |
| rs2747701 | FAM135A | 57579 | family with sequence similarity 135 member A |
| rs3732410 | GOLGB1 | 2804 | golgin B1 |
| rs31517 | LECT2 | 3950 | leukocyte cell derived chemotaxin 2 |
| rs242944 | SPPL2C | 162540 | signal peptide peptidase like 2C |
| rs9873604 | ZKSCAN7 | 55888 | zinc finger with KRAB and SCAN domains 7 |

**Figure 3:**
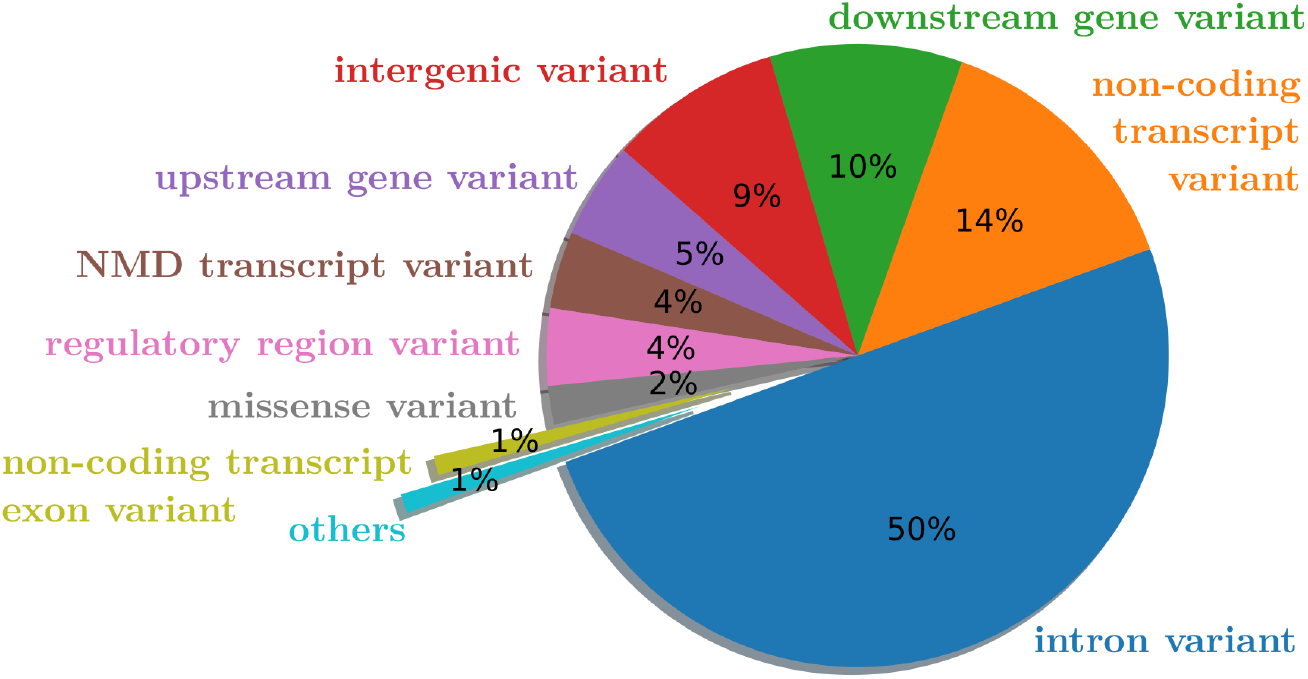
Functional consequences of our variants.

PheGenI results filtered 15/379 SNPs with significantly (*p* < 10^*−*5^) associated phenotypes, such as glaucoma, Behcet syndrome, autism spectrum disorder, stroke, among others, that may have a potential association with astigmatism. The sequence variant, rs3809863 located on chr17:4737646, was found to be significantly associated with the brain tissues, including the frontal cortex, hypothalamus, caudate basal ganglia in the appropriate effect size studies (*p* < 10^*−*8^). rs3809863 was found 379 bps apart from the cCRE, located within the genic region of ITGB3 and THCAT158. The *RegulomeDB* results also showed the intense transcription activity of this variant in the brain tissues, hippocampus, and substantia nigra. Similarly, other sequence variants were also found to be associated with brain disease-related phenotypes.

We also performed a variant-disease association study based on DisGeNET discovery platform [39]. We found 22 variants shown in Table 3 known to be annotated to various genetic disorders, such as depressive disorder, aortic aneurysm, bipolar disorder, hypertensive disease, alopecia, and so forth. Most of the diseases are either related to the astigmatism or co-exist with astigmatism. More details on these interlinks are illustrated in the subsequent sections.

#### 3.3.2 Astigmatism-related gene enrichment analysis

266 genes (say *G*_1_) of different types including but not limited to protein coding, pseudo gene, snRNA, or lincRNA - are harbored by our list of 379 variants *S*″. We have also compiled 181 genes (say *G*_2_) from DisGeNET database annotated with astigmatism with varied types (e.g. astigmatism, regular astigmatism - corneal, hyperopic astigmatism, etc). 5 genes (please, see Table 2) are common between *G*_1_ and *G*_2_ sets of genes. Overlap between these two groups of genes is statistically significant (*p* < 0.003). The statistical significance is measured using Fisher’s exact test by setting 49,000 genes as background. Here, background genes consist of protein coding genes, pseudo genes, genes expressing regulatory RNAs that do not encode proteins, and micro-RNA genes.

**Table 2:** Astigmatism-related 5 genes harbored by the variants identified by our methodology.

| Gene symbol | Gene name | Type |
| --- | --- | --- |
| CASC15 | cancer susceptibility 15 | lincRNA |
| EBF3 | EBF transcription factor 3 | protein coding |
| FBN1 | fibrillin 1 | protein coding |
| GRIN2B | glutamate ionotropic receptor NMDA type subunit 2B | protein coding |
| NFIX | nuclear factor I X | protein coding |

**Table 3:**
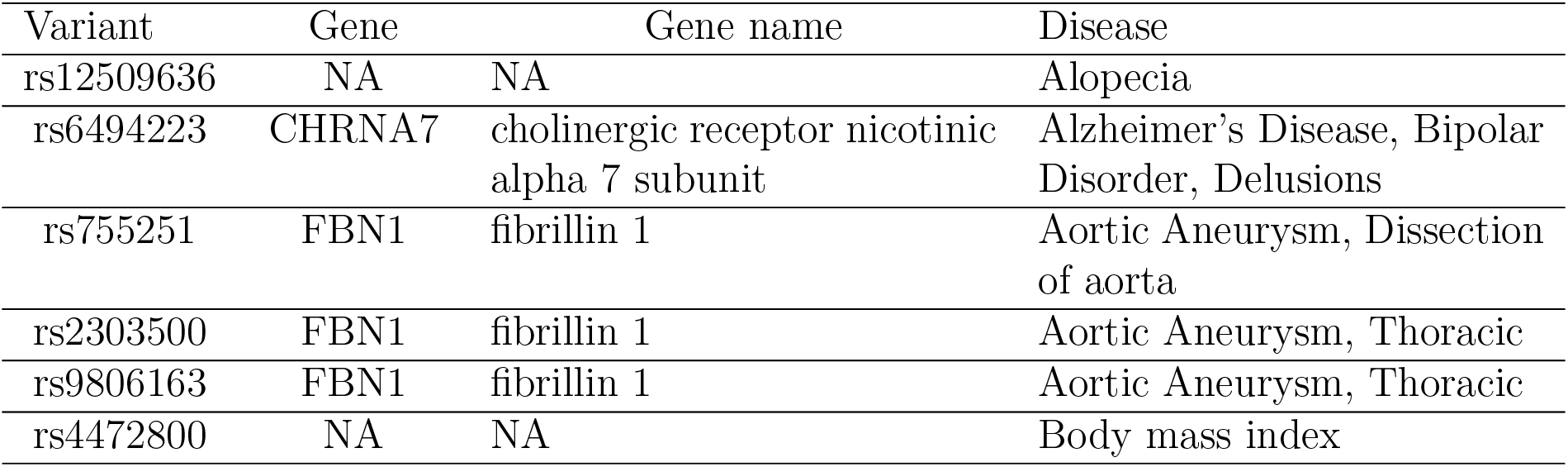
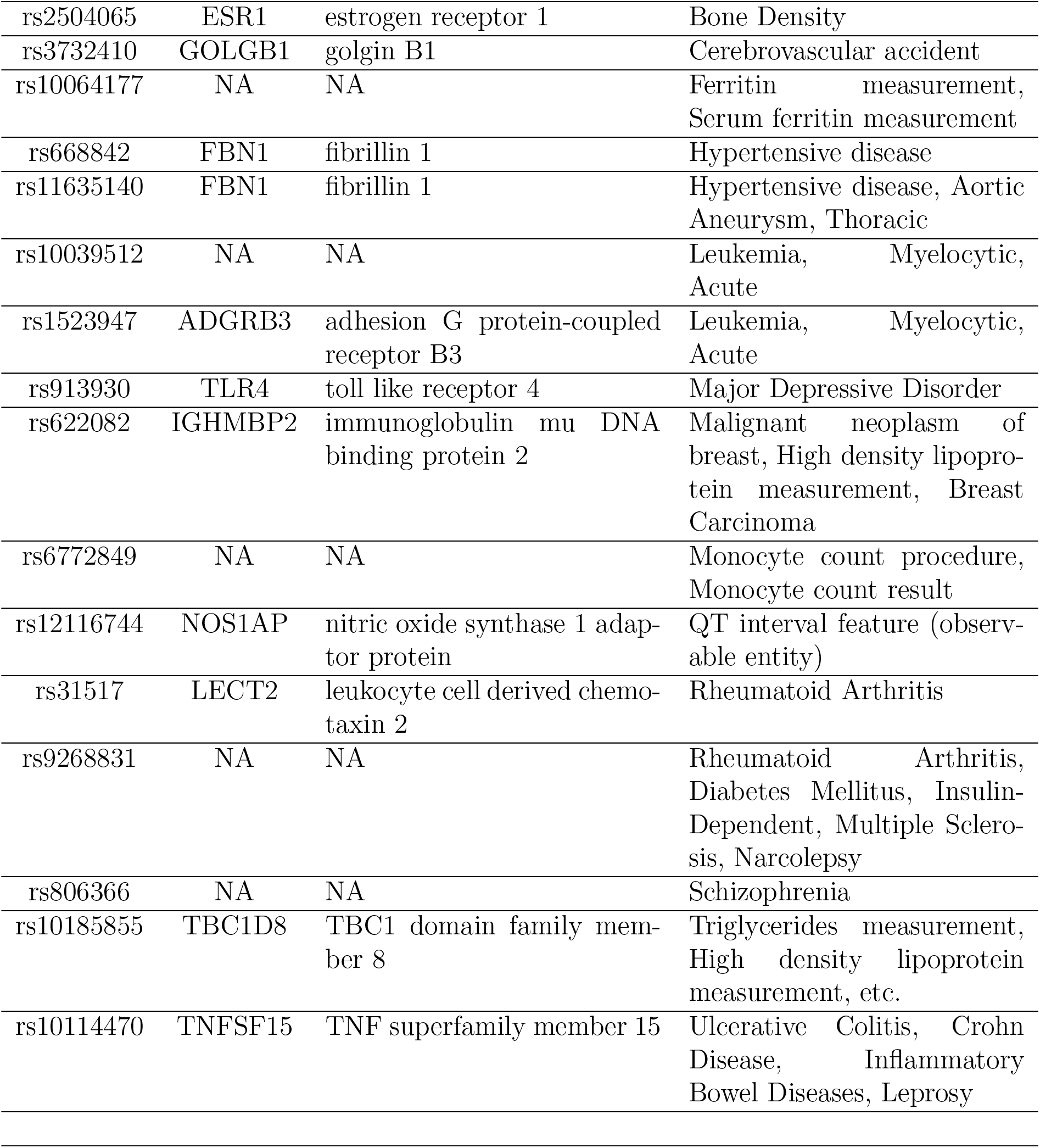
Variant-gene association based on DisNET.

| Variant | Gene | Gene name | Disease |
| --- | --- | --- | --- |
| rs12509636 | NA | NA | Alopecia |
| rs6494223 | CHRNA7 | cholinergic receptor nicotinic alpha 7 subunit | Alzheimer's Disease, Bipolar Disorder, Delusions |
| rs755251 | FBN1 | fibrillin 1 | Aortic Aneurysm, Dissection of aorta |
| rs2303500 | FBN1 | fibrillin 1 | Aortic Aneurysm, Thoracic |
| rs9806163 | FBN1 | fibrillin 1 | Aortic Aneurysm, Thoracic |
| rs4472800 | NA | NA | Body mass index |
| rs2504065 | ESR1 | estrogen receptor 1 | Bone Density |
| rs3732410 | GOLGB1 | golgin B1 | Cerebrovascular accident |
| rs10064177 | NA | NA | Ferritin measurement,<br>Serum ferritin measurement |
| rs668842 | FBN1 | fibrillin 1 | Hypertensive disease |
| rs11635140 | FBN1 | fibrillin 1 | Hypertensive disease, Aortic<br>Aneurysm, Thoracic |
| rs10039512 | NA | NA | Leukemia, Myelocytic,<br>Acute |
| rs1523947 | ADGRB3 | adhesion G protein-coupled<br>receptor B3 | Leukemia, Myelocytic,<br>Acute |
| rs913930 | TLR4 | toll like receptor 4 | Major Depressive Disorder |
| rs622082 | IGHMBP2 | immunoglobulin mu DNA<br>binding protein 2 | Malignant neoplasm of<br>breast, High density lipopro-<br>tein measurement, Breast<br>Carcinoma |
| rs6772849 | NA | NA | Monocyte count procedure,<br>Monocyte count result |
| rs12116744 | NOS1AP | nitric oxide synthase 1 adap-<br>tor protein | QT interval feature (observ-<br>able entity) |
| rs31517 | LECT2 | leukocyte cell derived chemo-<br>taxin 2 | Rheumatoid Arthritis |
| rs9268831 | NA | NA | Rheumatoid Arthritis,<br>Diabetes Mellitus, Insulin-Dependent, Multiple Sclero-<br>sis, Narcolepsy |
| rs806366 | NA | NA | Schizophrenia |
| rs10185855 | TBC1D8 | TBC1 domain family mem-<br>ber 8 | Triglycerides measurement,<br>High density lipoprotein<br>measurement, etc. |
| rs10114470 | TNFSF15 | TNF superfamily member 15 | Ulcerative Colitis, Crohn<br>Disease, Inflammatory<br>Bowel Diseases, Leprosy |

To perform the following studies, we consider only protein coding genes (say *G*_3_) contained by *G*_1_. The total number of such genes is 175 (i.e. |*G*_3_| = 175).

#### 3.3.3 Disease ontology (DO) term enrichment analysis

As stated above we performed DO (https://disease-ontology.org) term analysis by considering protein coding genes in *G*_3_. We found 8 Benjamini-Hochberg corrected (FDR < 0.05) DO terms as shown in Figure 4. Remarkably, most of the enriched terms are associated with mental health and autism spectrum disorder. According to Centers for Disease Control and Prevention (CDC), autism spectrum disorder (ASD) is a developmental disability that can cause serious social, communication, and behavioral challenges (https://www.cdc.gov/ncbddd/autism/facts.html). Various clinical studies suggest that there exists a strong correlation between ASD and astigmatism [40, 41]. Children with ASD have motor, sensory, language, and social-emotional delays that affect visual processing. Correspondingly, visual problems affect cognitive, speech-language, social-emotional, and perceptual development. Corneal astigmatism is significantly more frequent among children with ASD with respect to the typically developed population group (46.2% and 25.6%, respectively) [40]. Persons having mental disorder are often diagnosed with a diverse set of ocular problems, such as uncorrected refractive error, strabismus, blepharitis, pigmentary retinopathy, and cataracts [42]. Compound astigmatism was identified in a statistically significant number of individuals with mental illness [43].

**Figure 4:**
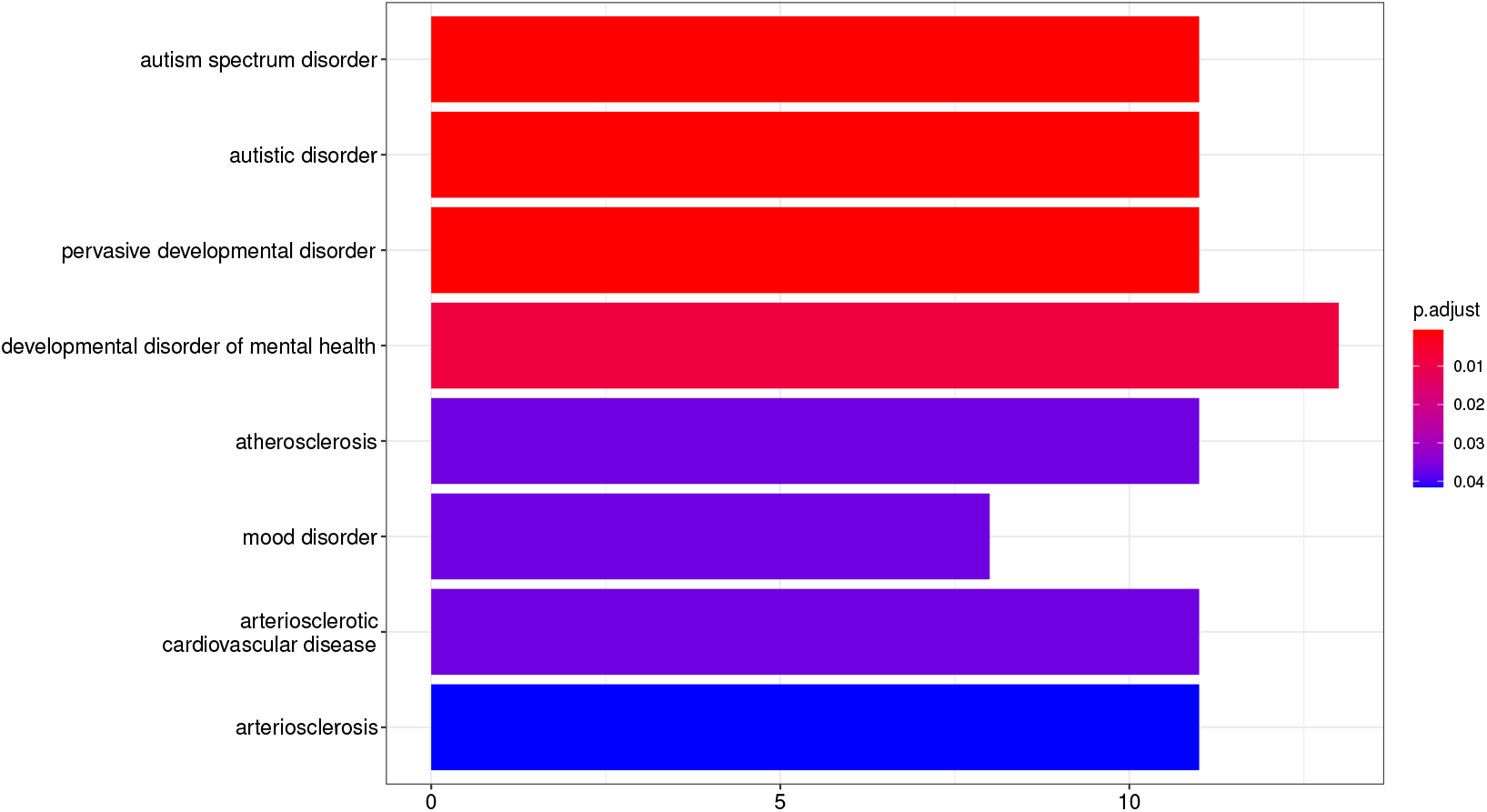
Benjamini-Hochberg corrected (FDR < 0.05) Disease Ontology (DO) terms.

We didn’t find any direct link between arteriosclerosis and astigmatism. But retinal disease is caused by arteriosclerosis. In this case, the arterioles (small arteries) in the retina are partly blocked due to the thickening of their walls. It is also responsible for cardiovascular disease. Individuals having cardiovascular disease may be at a greater possibility of developing certain types of eye related problems. According to the American Academy of Ophthalmology (AAO), research indicates that people suffering from heart disease have a higher chance of developing ophthalmic diseases (https://www.aao.org/eyenet/article/heart-eye-seeing-links). More can be found in [44].

#### 3.3.4 Pathway enrichment analysis

We found 14 Benjamini-Hochberg corrected (FDR < 0.05) enriched biological pathways from CPDB shown in Table 4. Please, note that while performing Hypergeometric over-representation test (https://en.wikipedia.org/wiki/Hypergeometric_distribution), we consider 21,500 as the number of background protein-coding genes. Several pathways are linked to ASD and ASD like diseases. For example, glutamatergic synapses consisting of glutamate localized inside presynaptic vesicles are the major excitatory synapses in our brain. Moreover, they have glutamate receptors on the postsynaptic membrane. Several clinical research suggest glutamate abnormalities in autism [45, 46, 47]. Moreover, Neurexins and neuroligins are synaptic cell-adhesion molecules.They connect pre- and postsynaptic neurons at synapses and define synaptic functions by mediating trans-synaptic signaling. In humans, alterations in neurexin or neuroligin genes are implicated in autism and other cognitive disorders [48, 49, 50]. NRXN1 (Neurexin 1) is one of the genes contained in Neurexins and neuroligins pathway. In a recent GWA study, 9 near genome-wide significant SNPs in NRXN1 gene were identified for refractive astigmatism [51]. In our study, our methodology identified rs3861561 SNP mutated in NRXN1 gene (i.e. this variant is one of the 350 discriminating variants identified by our algorithms). Moreover, we have found 4 genes enriched in Neurexins and neuroligins pathway as shown in Table 4.

**Table 4:** Benjamini-Hochberg corrected (FDR < 0.05) enriched biological pathways from different sources.

| Pathway | Source | FDR | Genes |
| --- | --- | --- | --- |
| Vitamin D-sensitive calcium signaling in depression | Wikipathways | 0.015796979 | ITPR1, BCL2, GRIN2B, SLC8A1 |
| Rap1 signaling pathway | KEGG | 0.015796979 | EFNA3, CNR1, ITGB3, MAGI2, RAPGEF5, GRIN2B, EFNA4, IGF1R] |
| Protein-protein interactions at synapses | Reactome | 0.02052683 | PTPRS, NRXN1, NRXN3, SHANK3, GRIN2B |
| SLC-mediated transmembrane transport | Reactome | 0.02052683 | SLC7A5, SLC25A26, SLC38A1, SLC14A2, SLCO3A1, SLC1A7, SLC8A1, SLC28A3 |
| Neurexins and neuroligins | Reactome | 0.021135656 | NRXN1, NRXN3, SHANK3, GRIN2B |
| Transport of inorganic cations/anions and amino acids/oligopeptides | Reactome | 0.022244722 | SLC7A5, SLC25A26, SLC38A1, SLC1A7, SLC8A1 |
| Disruption of post-synaptic signaling by CNV | Wikipathways | 0.027148897 | NRXN1, NRXN3, GRIN2B |
| Glutamatergic synapse | KEGG | 0.027148897 | SLC38A1, ITPR1, SLC1A7, GRIN2B, SHANK3 |
| Fragile X Syndrome | Wikipathways | 0.029042295 | CPT1A, CNR1, MAP1B, ITPR1, GRIN2B |
| Axon guidance | KEGG | 0.031736498 | SEMA5A, EFNA3, DCC, SEMA3A, EPHB1, EFNA4 |
| G alpha (12/13) signalling events | Reactome | 0.031736498 | TIAM2, ARHGEF3, ADRA1A, KALRN |
| Ebola Virus Pathway on Host | Wikipathways | 0.031736498 | NPC1, ITGB3, HLA-F, TLR4, IGF1R |
| Ion homeostasis | Reactome | 0.036534827 | ITPR1, SLC8A1, TRDN |
| Rett syndrome causing genes | Wikipathways | 0.043243418 | CECR2, SHANK3, GRIN2B |

Fragile X syndrome is another interesting pathway. Reportedly, it is the most frequently diagnosed inherited cause of intellectual disability. Fragile X syndrome co-occurs with autism in many cases and is a suspected genetic cause of autism in these cases. The prevalence of co-occurrence has been estimated to be 15% to 60% [52]. Moreover, a statistically significant number of individuals with Fragile X syndrome are associated with astigmatism [53, 54]. On the other hand, Rett syndrome is a rare genetic neurological and developmental disorder [55]. It affects the way the brain develops and eventually leads to loss of motor skills and speech progressively. Fragile X syndrome mainly affects boys where Rett disorder primarily affects girls [56].

One of the intriguing pathways found in our study is Ebola virus pathway. Ebola virus disease (EVD) is significantly associated with mental health consequences (such as, depression, anxiety, or PTSD) [57]. On the other hand, uveitis (a form of eye inflammation) is the most prevalent finding during the recovery from EVD [58]. Some of the ocular complications arising from uveitis consist of cataract, retinal scarring, optic neuropathy, hypotony, and phthisis bulbi. Consequently, they can lead to severe vision impairment or even complete bilateral blindness of the affected individuals.

#### 3.3.5 Pathway network analysis

We performed pathway network analysis to find functional modules, pathway significance, and gene clusters based on graph theory as discussed in [59]. The method works as follows: It builds a weighted network based on the enriched pathways. Each pathway represents a node in the graph and each pair of nodes is connected via an edge if-and-only-if they have some common genes. The weight of the edge between any pairs of nodes is the similarity between that particular pair. Weight of an edge is calculated based on the common GO-BP terms possessed by the genes in the pathways (i.e. nodes) incident on that edge. After building the weighted network, we can compute the importance score of each pathway using the graph centrality measure (such as, closeness centrality, betweenness centrality, etc.). Moreover, we can disassemble the network into a set of functional modules based on any suitable graph clustering algorithms (such as spectral clustering, Louvain clustering, etc.). It is evident from the article [59] that the method accurately identifies functional modules mimicking real biological events.

Based on the set of protein coding genes *G*_3_, we found 14 enriched pathways as described above. We build a weighted network based on those 14 pathways using the method described above. The corresponding weighted network is shown in Figure 8. The diameter of a node is proportional to its importance. Similarly, the width of an edge is proportional to its weight (i.e. similarity between two incident nodes of this edge). It is noticeable from the figure that *Neurexins and neuroligins* and *Ebola Virus Pathway on Host* is the most and the least important pathway in this network, respectively. We find two distinct functional modules in this network, e.g. (1) developmental disorder and synaptic activity related module (*C*1) consisting of 10 pathways and (2) transport related module (*C*2) comprising of 4 pathways. Correspondingly, we found 2 gene clusters; each gene cluster contained by each of the functional modules. Gene cluster for *C*1 and *C*2 contains 29 genes and 12 genes (out of 175 protein coding genes *G*_3_), respectively as shown in Figure 9. 4 genes are common in those 2 gene clusters, namely GRIN2B (Glutamate Ionotropic Receptor NMDA Type Subunit 2B), ITPR1 (Inositol 1,4,5-trisphosphate receptor type 1), SLC1A7 (Solute Carrier Family 1 Member 7), and SLC38A1 (Solute Carrier Family 38 Member 1).

#### 3.3.6 Gene ontology (GO) term enrichment analysis

We performed GO-BP, GO-MF, and GO-CC (http://geneontology.org) term enrichment analyses based on the protein coding genes *G*_3_ found by our methodology. We retained only enriched terms having FDR < 0.05. We found 58 enriched GO-BP terms and top 20 such terms are displayed in Figure 5. Likewise, we obtained 26 enriched GO-CC terms and displayed the top 20 in Figure 6. In addition, Figure 7 shows all the enriched 11 GO-MF terms. As we can see, most of the terms are associated with various aspects of synapses, synaptic activity, and functions. According to this excellent review [50], pathogenesis of ASD may, at least in part, be attributed to synaptic dysfunction.

**Figure 5:**
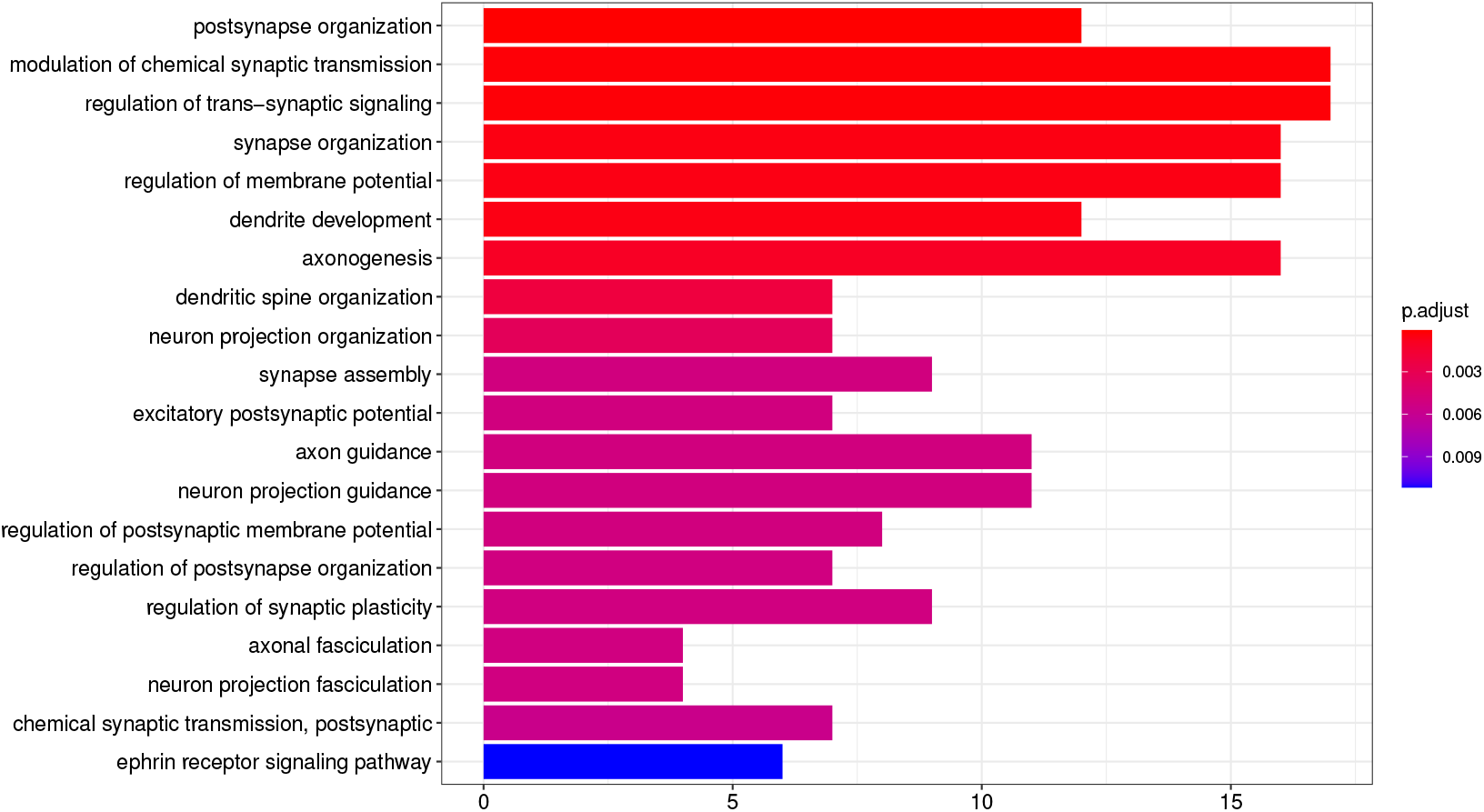
Top 20 enriched (FDR < 0.05) Gene Ontology-Biological Process (GO-BP) terms.

**Figure 6:**
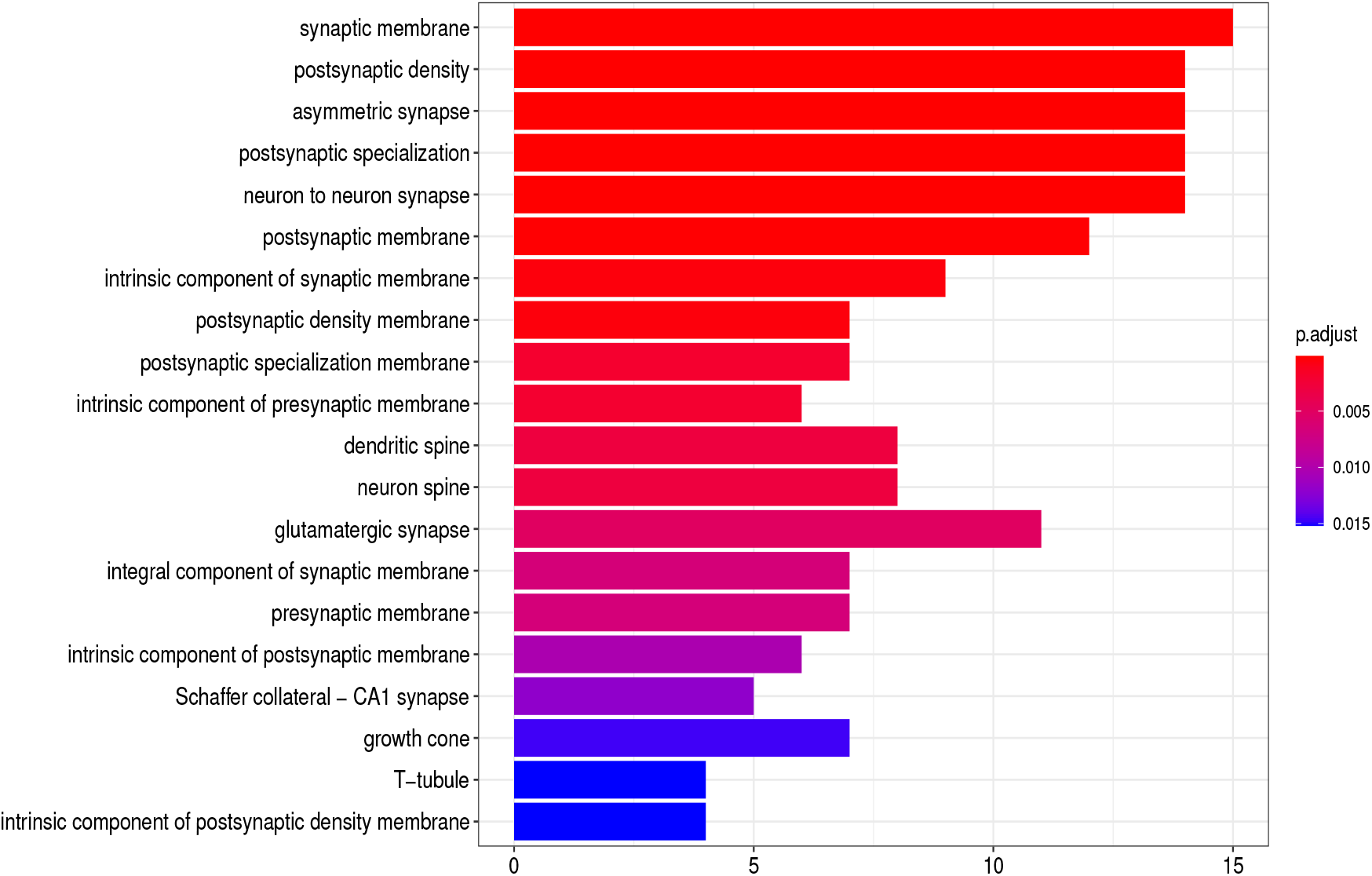
Top 20 enriched (FDR < 0.05) Gene Ontology-Cellular Component (GO-CC) terms.

**Figure 7:**
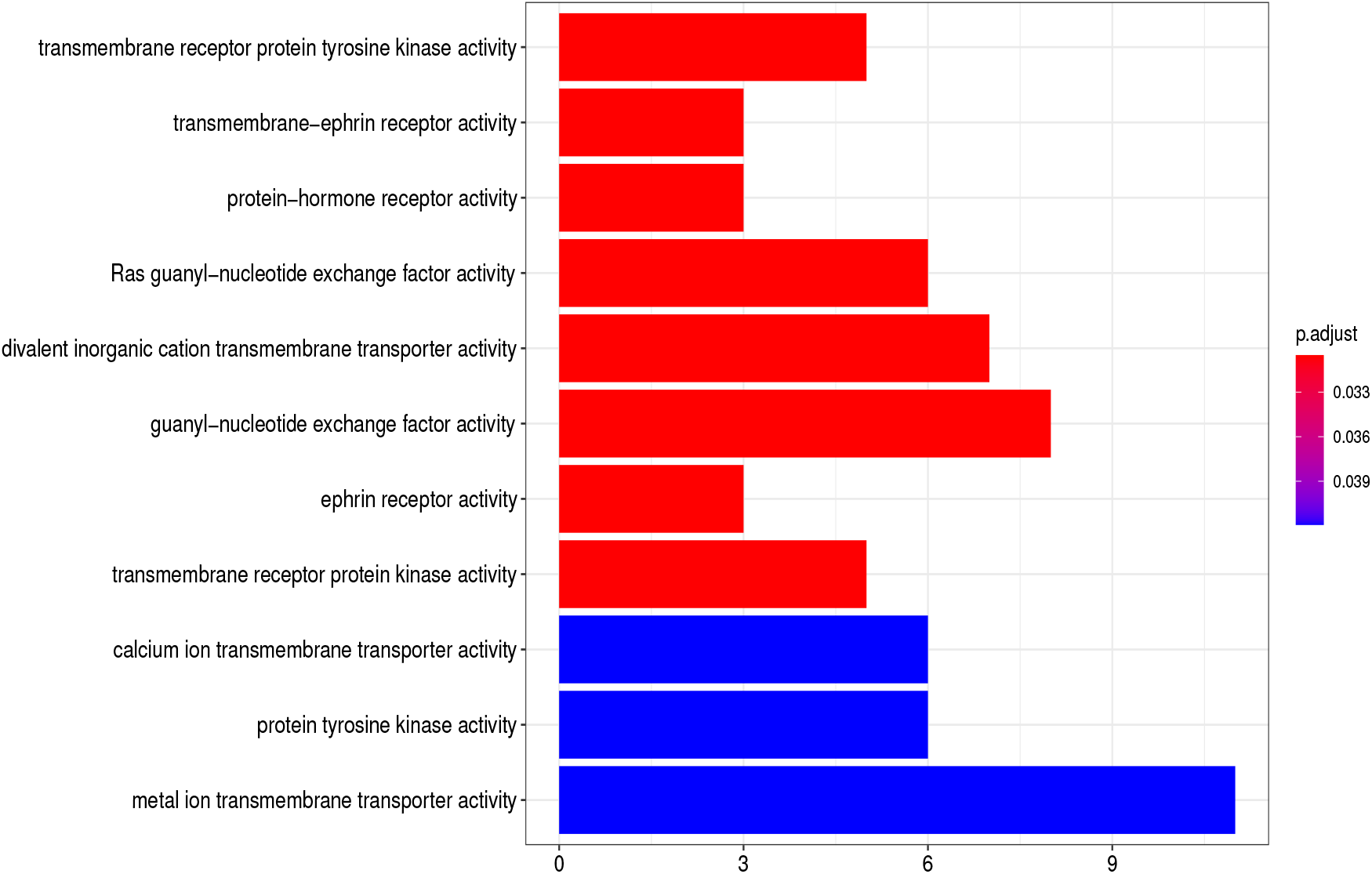
Enriched (FDR < 0.05) Gene Ontology-Molecular Function (GO-MF) terms.

**Figure 8:**
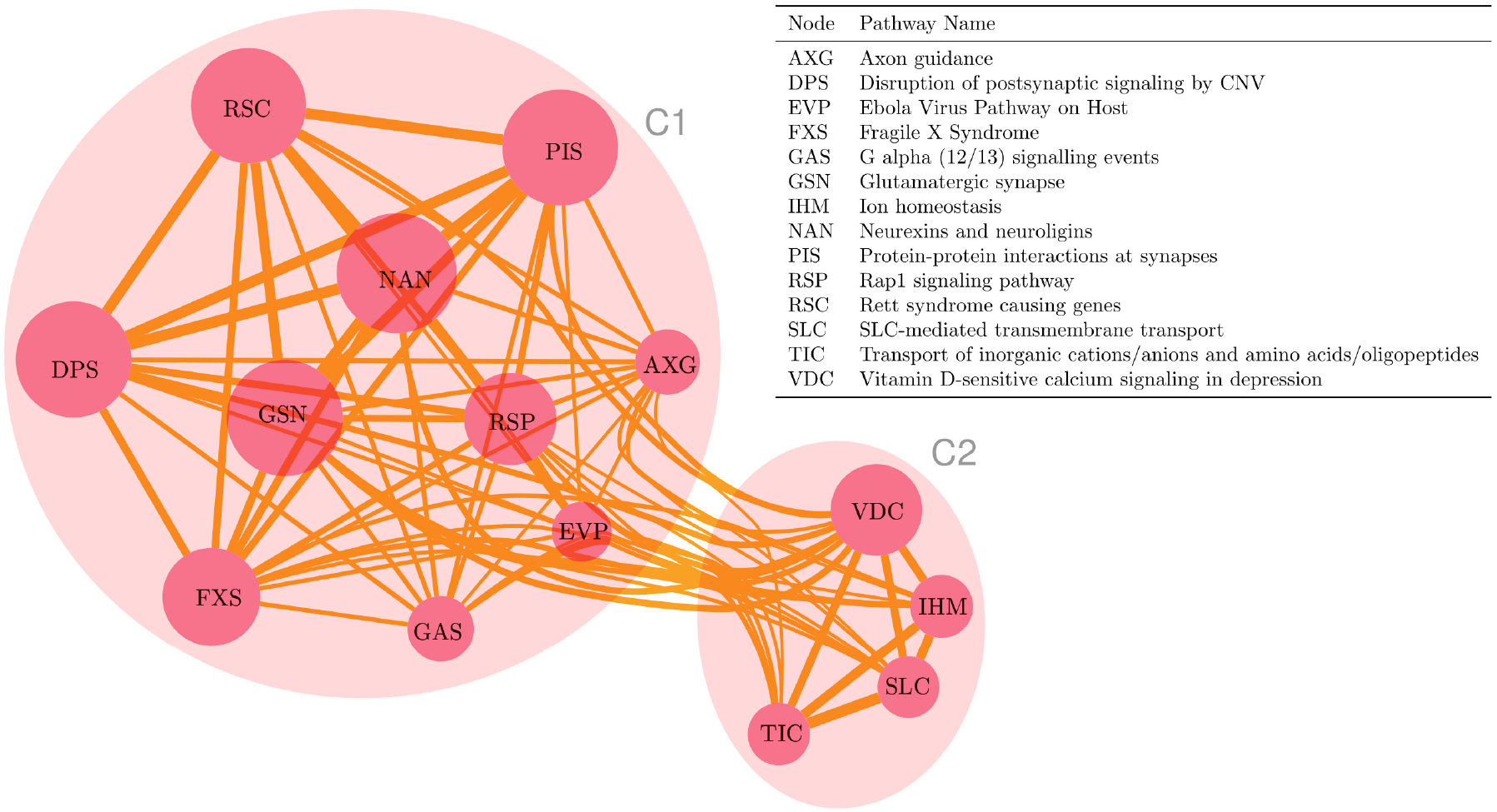
Network of enriched (FDR < 0.05) biological pathways.

**Figure 9:**
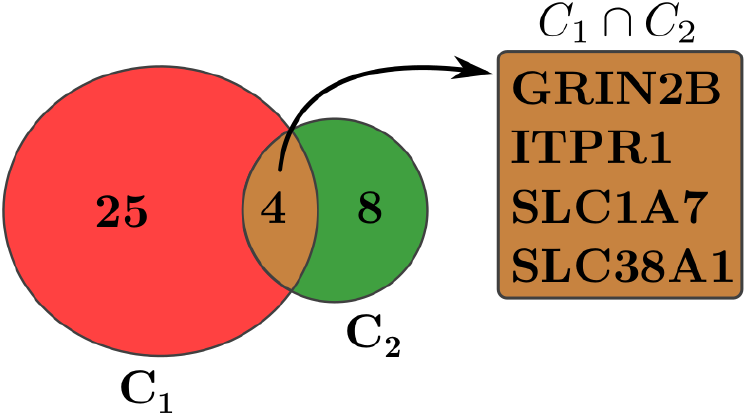
Gene clusters found in the pathway network.

## 4 Discussions

In this section we discuss two topics categorically. These are as follows:

### 4.1 Our proposed methodology

There is a lack of reliable machine learning methodology for solving complex multi-locus problems in GWAS. The problem is inherently very hard to solve as the subset selection is known to be NP-hard and thus, it is not solvable in polynomial time in current technology (https://en.wikipedia.org/wiki/NP-hardness). Consequently, we don’t have any choices but to accept approximate solutions. The approximate solution should be context specific and near optimal. I.e., in the medical domain the solution should elucidate the underlying convoluted biology. In a typical GWAS, we have several hundreds of thousands to millions of variants. On the contrary, the number of samples is not more than a few hundreds to a handful of thousands. As the system is very underdetermined, no unique solution exists. Consequently, we will have several solutions with similar significance and even worse, some solution may be spurious. In addition to this, with a slight change in the dataset, we will likely get a very different solution.

To address the issues stated above, we propose a reliable learning algorithm that can identify a stable and robust subset of discriminating variants. Stability is ensured by slightly decreasing the number of samples multiple times. Robustness is ensured by bootstrapping the samples multiple times. The observation is that the variants that are genuinely discriminating should prevail under slight changes in the dataset. The spurious ones will be ousted eventually. In the domain of machine learning, ensemble learning refers to multiple machine learning algorithms being used to search multiple hypothesis spaces and produce good approximations to the true ones. But the way we formulate it is novel. At first, we find a set of lists containing discriminating variants having the highest classification accuracy from bootstrapped samples. Then we return the final set of discriminating variants based on their stability, robustness, and classification accuracy across the lists.

*The curse of dimensionality* is another problem of solving multi-locus problems in GWAS. We should avoid *p*-value thresholding as much as possible. We have addressed this issue by discarding a set of variants with a little loss of information content in the GWAS dataset. The framework discards all the duplicates at the beginning based on randomization and graph theory. An intuitive LD pruning algorithm based on graph theory is devised to select each tagged variant based on its PageRank centrality score in the entire network within a window of size 400kb.

### 4.2 Biology of astigmatism

The cause of astigmatism is complicated and not fully explained. Environmental effects, such as the number of hours engaging in video games was found to be correlated with more severe astigmatism. This study was conducted on school going 7-9 years old children. However, other studies found a significant genetic contribution to astigmatism, such as the risk of developing astigmatism is 2-*fold* in first degree relatives of persons having astigmatism. Moreover, family and twin studies indicate a roughly 60% heritability of astigmatism [60, 61, 62].

We have identified a set of discriminating variants as discussed in the result section. Next, we discuss one of them in detail. The sequence variant, rs3809863, is associated with astigmatism and open-angle glaucoma [63]. Additionally, the variant, located within the genic region of the ITGB3 and THCAT158 (also known as EFCAB13-DT: EFCAB13 divergent transcript), was found to be near to the cCREs showing a high association with the transcription. ITGB3 was found to be deregulated and may be involved in intraocular pressure elevation. Elevation of intraocular pressure may further lead to the destruction of the retinal structure during the glaucoma progression [64]. In addition to this, myopia was also observed to be associated with glaucoma [65]. Moreover, myopia was also reported to be related to astigmatism. However, a direct relation is still needed to be established [66]. The variant was also located into the genic region of the THCAT158 which is a non-coding RNA (ncRNA). Although the role of the gene is not known but ncRNAs were found to be associated with myopia as well as irregular astigmatisms [67]. Therefore, we believe that the THCAT158 may be associated with the astigmatism which needs to be investigated further. Moreover, glaucoma caused by progressive optic nerve degeneration is considered a neurodegenerative disorder of both the eye and the brain neurodegeneration [68]. Although there is no good consensus regarding the exact association of astigmatism with the above clinical manifestations, variants identified in astigmatism and their association with glaucoma, brain tissue enrichment, and other neurodegenerative phenotypes suggest a link with astigmatism.

Current studies (as discussed in Results section) suggest that autism, mental illness, or mental retardation are directly connected with astigmatism. In this investigation, we have also established similar facts and observations. There is no known causal effect of autism or any mental disorders to astigmatism and vice-versa. Consequently, a significant number of dual diagnoses of astigmatism and autism/autism like diseases suggest that both likely share some common molecular pathways. It also suggests that there should be a significant amount of genetics involved in astigmatism. More research should be done to elucidate this disorder. If we can fully understand the common genetics behind the co-occurrence of astigmatism and various developmental genetic disorders, astigmatism can be used as a pre-screening tool for early detection and intervention of such illnesses.

## 5 Conclusions

In this article we have proposed a thorough methodology to solve multi-locus problems in GWAS based on graph theory and machine learning. At first, we efficiently identify all the duplicate variants and retain only unique ones based on randomization and graph theory. Next, we select representative variants and eliminate the rest having high LDs with those representatives based on an intuitive graph theoretic algorithm. Finally, we offer a robust and stable machine learning algorithm for identifying a subset of discriminating variants that can effectively explain the genetics behind a complex disease. We have performed rigorous experiments by taking a GWAS case-control dataset where cases are persons having astigmatism and controls do not have any astigmatisms. The results indicate that our proposed framework is indeed able to decipher the genetics of astigmatism from such a small set of samples where classical GWAS fails to identify any genome-wide significant variants.

## Data Availability

All data produced in the present study are available upon reasonable request to the authors.

## References

[1] Balding, D.: A tutorial on statistical methods for population association studies. Nature Reviews Genetics 7, 781–791 (2006)

[2] Cordell, H.: Epistasis: what it means, what it doesn’t mean, and statistical methods to detect it in humans. Human molecular genetics 11 20, 2463–8 (2002)

[3] Gaudillo, J., Rodriguez, J.J.R., Nazareno, A., Baltazar, L.R.P., Vilela, J., Bulalacao, R., Domingo, M., Albia, J.: Machine learning approach to single nucleotide polymorphism-based asthma prediction. PLoS ONE 14 (2019)

[4] Mieth, B., Rozier, A., Rodriguez, J.A., Höhne, M.M., Görnitz, N., Müller, K.-R.: Deepcombi: explainable artificial intelligence for the analysis and discovery in genome-wide association studies. NAR Genomics and Bioinformatics 3 (2021)

[5] Xu, M., Tantisira, K., Wu, A., Litonjua, A., Chu, J.-h., Himes, B., Damask, A., Weiss, S.: Genome wide association study to predict severe asthma exacerbations in children using random forests classifiers. BMC Medical Genetics 12, 90–90 (2010)

[6] Mieth, B., Kloft, M., Rodriguez, J.A., Sonnenburg, S., Vobruba, R., Morcillo-Suarez, C., Farré, X., Marigorta, U., Fehr, E., Dickhaus, T., Blanchard, G., Schunk, D., Navarro, A., Müller, K.-R.: Combining multiple hypothesis testing with machine learning increases the statistical power of genome-wide association studies. Scientific Reports 6 (2016)

[7] Listgarten, J., Damaraju, S., Poulin, B., Cook, L., Dufour, J., Driga, A., Mackey, J., Wishart, D., Greiner, R., Zanke, B.: Predictive models for breast cancer susceptibility from multiple single nucleotide polymorphisms. Clinical Cancer Research 10, 2725–2737 (2004)

[8] Hajiloo, M., Damavandi, B., HooshSadat, M., Sangi, F., Mackey, J., Cass, C., Greiner, R., Damaraju, S.: Breast cancer prediction using genome wide single nucleotide polymorphism data. BMC Bioinformatics 14, 3–3 (2013)

[9] Romagnoni, A., Jégou, S., Steen, K.V., Wainrib, G., Hugot, J., Peyrin-Biroulet, L., et al.: Comparative performances of machine learning methods for classifying crohn disease patients using genome-wide genotyping data

[10] Bolón-Canedo, V., Alonso-Betanzos, A.: Ensembles for feature selection: A review and future trends. Inf. Fusion 52, 1–12 (2019)

[11] Slatkin, M.: Linkage disequilibrium — understanding the evolutionary past and mapping the medical future. Nature Reviews Genetics 9, 477–485 (2008)

[12] Hill, W., Robertson, A.: Linkage disequilibrium in finite populations. Theoretical and Applied Genetics 38, 226–231 (2005)

[13] Sullivan, D.: What Is Google PageRank? A Guide For Searchers & Webmasters. https://searchengineland.com/what-is-google-pagerank-a-guide-for-searchers-webmasters-11068 Accessed Accessed 1 Sep 2021

[14] Boser, B., Guyon, I., Vapnik, V.: A training algorithm for optimal margin classifiers. In: COLT ‘92 (1992)

[15] Cortes, C., Vapnik, V.: Support-vector networks. Machine Learning 20, 273–297 (1995)

[16] Müller, K., Mika, S., Rätsch, G., Tsuda, K., Schölkopf, B.: An introduction to kernel-based learning algorithms. IEEE transactions on neural networks 12 2, 181–201 (2001)

[17] Joachims, T.: Training linear svms in linear time. In: Proceedings of the 12th ACM SIGKDD International Conference on Knowledge Discovery and Data Mining - KDD ‘06, p. 217. ACM Press, ??? (2006). doi:10.1145/1150402.1150429. http://portal.acm.org/citation.cfm?doid=1150402.1150429

[18] 1. Howe, K.L., Achuthan, P., Allen, J., Allen, J., Alvarez-Jarreta, J., Amode, M.R., Armean, I.M., Azov, A.G., Bennett, R., Bhai, J., et al.: Ensembl 2021. Nucleic Acids Research 49(D1), 884–891 (2021). doi:10.1093/nar/gkaa942

[19] Ng, P.C.: Sift: predicting amino acid changes that affect protein function. Nucleic Acids Research 31(13), 3812–3814 (2003). doi:10.1093/nar/gkg509

[20] Ramensky, V.: Human non-synonymous snps: server and survey. Nucleic Acids Research 30(17), 3894–3900 (2002). doi:10.1093/nar/gkf493

[21] Rentzsch, P., Witten, D., Cooper, G.M., Shendure, J., Kircher, M.: Cadd: predicting the deleteriousness of variants throughout the human genome. Nucleic Acids Research 47(D1), 886–894 (2019). doi:10.1093/nar/gky1016

[22] Ioannidis, N.M., Rothstein, J.H., Pejaver, V., Middha, S., McDonnell, S.K., Baheti, S., Musolf, A., Li, Q., Holzinger, E., Karyadi, D., et al.: Revel: An ensemble method for predicting the pathogenicity of rare missense variants. The American Journal of Human Genetics 99(4), 877–885 (2016). doi:10.1016/j.ajhg.2016.08.016

[23] Ramos, E.M., Hoffman, D., Junkins, H.A., Maglott, D., Phan, L., Sherry, S.T., Feolo, M., Hindorff, L.A.: Phenotype–genotype integrator (phegeni): synthesizing genome-wide association study (gwas) data with existing genomic resources. European Journal of Human Genetics 22(1), 144–147 (2014). doi:10.1038/ejhg.2013.96

[24] Reva, B., Antipin, Y., Sander, C.: Predicting the functional impact of protein mutations: application to cancer genomics. Nucleic Acids Research 39(17), 118–118 (2011). doi:10.1093/nar/gkr407

[25] Hinrichs, A.S., Karolchik, D., Baertsch, R., Barber, G.P., Bejerano, G., Clawson, H., Diekhans, M., Furey, T.S., Harte, R.A., Hsu, F., et al.: The ucsc genome browser database: update 2006. Nucleic Acids Research 34(suppl 1), 590–598 (2006). doi:10.1093/nar/gkj144

[26] Quinlan, A.R., Hall, I.M.: Bedtools: a flexible suite of utilities for comparing genomic features. Bioinformatics 26(6), 841–842 (2010). doi:10.1093/bioinformatics/btq033

[27] Boyle, A.P., Hong, E.L., Hariharan, M., Cheng, Y., Schaub, M.A., Kasowski, M., Karczewski, K.J., Park, J., Hitz, B.C., Weng, S., et al.: Annotation of functional variation in personal genomes using regulomedb. Genome Research 22(9), 1790–1797 (2012). doi:10.1101/gr.137323.112

[28] Tzovaras, B.G., Rausch, H., Bayer, P.: openSNP. https://opensnp.org/ Accessed Accessed 1 Sep 2021

[29] Gustavsen, J., Rüeger, S., Chamberlain, S., Ushey, K., Zhu, H.: Rsnps: Get ‘SNP’ (‘Single-Nucleotide’ ‘Polymorphism’) Data on the Web. (2020). R package version 0.4.0. https://CRAN.R-project.org/package=rsnps

[30] Purcell, S.: PLINK 1.9. https://zzz.bwh.harvard.edu/plink/ Accessed Accessed 1 Sep 2021

[31] Purcell, S., Neale, B., Todd-Brown, K., Thomas, L., Ferreira, M.A., Bender, D., Maller, J., Sklar, P., De Bakker, P.I., Daly, M.J., et al.: Plink: a tool set for whole-genome association and population-based linkage analyses. The American journal of human genetics 81(3), 559–575 (2007)

[32] Marees, A., de Kluiver, H., Stringer, S., Vorspan, F., Curis, E., Marie-Claire, C., Derks, E.: A tutorial on conducting genome-wide association studies: Quality control and statistical analysis. International Journal of Methods in Psychiatric Research 27 (2018)

[33] Wu, T., Hu, E., Xu, S., Chen, M., Guo, P., Dai, Z., Feng, T., Zhou, L., Tang, W., Zhan, L., Fu, X., Liu, S., Bo, X., Yu, G.: clusterprofiler 4.0: A universal enrichment tool for interpreting omics data. The Innovation 2(3), 100141 (2021). doi:10.1016/j.xinn.2021.100141

[34] Kamburov, A., Wierling, C., Lehrach, H., Herwig, R.: Consensuspathdb—a database for integrating human functional interaction networks. Nucleic Acids Research 37, 623–628 (2009)

[35] McLaren, W., Gil, L., Hunt, S., Riat, H., Ritchie, G., Thormann, A., Flicek, P., Cunningham, F.: The ensembl variant effect predictor. Genome Biology 17 (2016)

[36] Read, S.A., Collins, M.J., Carney, L.G.: A review of astigmatism and its possible genesis. Clinical and Experimental Optometry 90(1), 5–19 (2007). doi:10.1111/j.1444-0938.2007.00112.x. https://doi.org/10.1111/j.1444-0938.2007.00112.x

[37] Lopes, M.C., Hysi, P., Verhoeven, V., MacGregor, S., Hewitt, A., Montgomery, G., Cumberland, P., Vingerling, J., Young, T., van Duijn, C.V., Oostra, B., Uitterlinden, A., Rahi, J., Mackey, D., Klaver, C., Andrew, T., Hammond, C.: Identification of a candidate gene for astigmatism. Investigative ophthalmology & visual science 54 2, 1260–7 (2013)

[38] Mozayan, E., Lee, J.K.: Update on astigmatism management. Current Opinion in Ophthalmology 25, 286–290 (2014)

[39] González, J., Bravo, Á., Queralt-Rosinach, N., Gutiérrez-Sacristán, A., Deu-Pons, J., Centeno, E., Garćıa-Garćıa, J., Sanz, F., Furlong, L.: Disgenet: a comprehensive platform integrating information on human disease-associated genes and variants. Nucleic Acids Research 45, 833–839 (2017)

[40] Anketell, P., Saunders, K., Gallagher, S., Bailey, C., Little, J.: Profile of refractive errors in european caucasian children with autistic spectrum disorder; increased prevalence and magnitude of astigmatism. Ophthalmic and Physiological Optics 36, 395–403 (2016)

[41] Ikeda, J., Davitt, B., Ultmann, M., Maxim, R., Cruz, O.: Brief report: Incidence of ophthalmologic disorders in children with autism. Journal of Autism and Developmental Disorders 43, 1447–1451 (2013)

[42] Maino, D., Rado, M., Pizzi, W.: Ocular anomalies of individuals with mental illness and dual diagnosis. Journal of the American Optometric Association 67 12, 740–8 (1996)

[43] Harvey, E., McGrath, E., Miller, J.M., Davis, A.L., Twelker, J.D., Dennis, L.: A preliminary study of astigmatism and early childhood development. Journal of AAPOS: the official publication of the American Association for Pediatric Ophthalmology and Strabismus 22 4, 294–298 (2018)

[44] Cheung, C., Wong, T.: Is age-related macular degeneration a manifestation of systemic disease? new prospects for early intervention and treatment. Journal of Internal Medicine 276, 140–153 (2014)

[45] Rojas, D.: The role of glutamate and its receptors in autism and the use of glutamate receptor antagonists in treatment. Journal of Neural Transmission 121, 891–905 (2014)

[46] Horder, J., Petrinovic, M., Mendez, M., Bruns, A., Takumi, T., Spooren, W., Barker, G., Künnecke, B., Murphy, D.: Glutamate and gaba in autism spectrum disorder—a translational magnetic resonance spectroscopy study in man and rodent models. Translational Psychiatry 8 (2018)

[47] Elst, L.V., Maier, S., Fangmeier, T., Endres, D., Mueller, G., Nickel, K., Ebert, D., Lange, T., Hennig, J., Biscaldi, M., Riedel, A., Perlov, E.: Disturbed cingulate glutamate metabolism in adults with high-functioning autism spectrum disorder: evidence in support of the excitatory/inhibitory imbalance hypothesis. Molecular Psychiatry 19, 1314–1325 (2014)

[48] Südhof, T.: Neuroligins and neurexins link synaptic function to cognitive disease. Nature 455, 903–911 (2008)

[49] Reichelt, A., Dachtler, J.: The role of neurexins and neuroligins in autism. (2015)

[50] Guang, S., Pang, N., Deng, X., Yang, L.-f., He, F., Wu, L., Chen, C., Yin, F., Peng, J.: Synaptopathology involved in autism spectrum disorder. Frontiers in Cellular Neuroscience 12 (2018)

[51] Li, Q., Wojciechowski, R., Simpson, C., Hysi, P., Verhoeven, V., Ikram, M., et al.: Genome-wide association study for refractive astigmatism reveals genetic codetermination with spherical equivalent refractive error: the cream consortium. Human Genetics 134, 131–146 (2014)

[52] Kaufmann, W., Kidd, S., Andrews, H., Budimirovic, D., Esler, A.N., Haas-Givler, B., Stackhouse, T., Riley, C., Peacock, G., Sherman, S., Brown, W., Berry-Kravis, E.: Autism spectrum disorder in fragile x syndrome: Cooccurring conditions and current treatment. Pediatrics 139, 194–206 (2017)

[53] Maino, D., Wesson, M., Schlange, D., Cibis, G., Maino, J.: Optometric findings in the fragile x syndrome. Optometry and vision science: official publication of the American Academy of Optometry 68 8, 634–40 (1991)

[54] Hatton, D., Buckley, E., Lachiewicz, A., Roberts, J.: Ocular status of boys with fragile x syndrome: a prospective study. Journal of AAPOS: the official publication of the American Association for Pediatric Ophthalmology and Strabismus 2 5, 298–302 (1998)

[55] Rett syndrome: MedlinePlus Genetics. https://medlineplus.gov/genetics/condition/rett-syndrome/ Accessed Accessed 1 Sep 2021

[56] Rett Syndrome Fact Sheet — National Institute of Neurological Disorders and Stroke. https://www.ninds.nih.gov/Disorders/Patient-Caregiver-Education/Fact-Sheets/Rett-Syndrome-Fact-Sheet Accessed Accessed 1 Sep 2021

[57] Shantha, J., Crozier, I., Yeh, S.: An update on ocular complications of ebola virus disease. Current Opinion in Ophthalmology 28, 600–606 (2017)

[58] Cénat, J., Felix, N., Blais-Rochette, C., Rousseau, C., Bukaka, J., Derivois, D., Noorishad, P.-G., Birangui, J.-P.: Prevalence of mental health problems in populations affected by the ebola virus disease: A systematic review and meta-analysis. Psychiatry Research 289 (2020)

[59] Saha, S., Soliman, A., Rajasekaran, S.: A novel pathway network analytics method based on graph theory. Journal of computational biology: a journal of computational molecular cell biology (2021)

[60] Grjibovski, A., Magnus, P., Midelfart, A., Harris, J.R.: Epidemiology and heritability of astigmatism in norwegian twins: An analysis of self-reported data. Ophthalmic Epidemiology 13, 245–252 (2006)

[61] Hammond, C., Snieder, H., Gilbert, C., Spector, T.: Genes and environment in refractive error: the twin eye study. Investigative ophthalmology & visual science 42 6, 1232–6 (2001)

[62] Dirani, M., Islam, A., Shekar, S., Baird, P.: Dominant genetic effects on corneal astigmatism: the genes in myopia (gem) twin study. Investigative ophthalmology & visual science 49 4, 1339–44 (2008)

[63] Vishal, M., Sharma, A., Kaurani, L., Alfano, G., Mookherjee, S., Narta, K., Agrawal, J., Bhattacharya, I., Roychoudhury, S., Ray, J., et al.: Genetic association and stress mediated down-regulation in trabecular meshwork implicates mpp7 as a novel candidate gene in primary open angle glaucoma. BMC Medical Genomics 9(1), 15 (2016). doi:10.1186/s12920-016-0177-6

[64] Rocha, L.R., Nguyen Huu, V.A., Palomino La Torre, C., Xu, Q., Jabari, M., Krawczyk, M., Weinreb, R.N., Skowronska-Krawczyk, D.: Early removal of senescent cells protects retinal ganglion cells loss in experimental ocular hypertension. Aging Cell 19(2) (2020). doi:10.1111/acel.13089

[65] Chen, S.-J., Lu, P., Zhang, W.-F., Lu, J.-H.: High myopia as a risk factor in primary open angle glaucoma. International Journal of Ophthalmology 5(6), 750–753

[66] Fulton, A.B., Hansen, R.M., Petersen, R.A.: The relation of myopia and astigmatism in developing eyes. Ophthalmology 89(4), 298–302 (1982). doi:10.1016/S0161-6420(82)34788-0

[67] Khaled, M.L., Bykhovskaya, Y., Yablonski, S.E.R., Li, H., Drewry, M.D., Aboobakar, I.F., Estes, A., Gao, X.R., Stamer, W.D., Xu, H., et al.: Differential expression of coding and long noncoding rnas in keratoconus-affected corneas. Investigative Opthalmology & Visual Science 59(7), 2717 (2018). doi:10.1167/iovs.18-24267

[68] Chan, J.W., Chan, N.C., Sadun, A.A.: ¡p¿glaucoma as neurodegeneration in the brain¡/p¿. Eye and Brain 13, 21–28 (2021). doi:10.2147/EB.S293765

